# Seroprevalence and predictors of Hepatitis A virus immunity among young MSM in urban Brazil: a cross-sectional study at a referral center

**DOI:** 10.1101/2025.08.28.25334666

**Authors:** Alberto dos Santos de Lemos, Marcellus Dias da Costa, Luciana Gomes Pedro Brandão, Sergio Carlos Assis de Jesus, Daniel Marinho da Costa, Ananza Taina da Silva Villaça, Flavio de Carvalho, Margareth Catoia Varela, Pedro Emmanuel Alvarenga Americano do Brasil

## Abstract

In addition, the usual socioeconomic and sanitary exposures to Hepatitis A Virus (HAV), men who have sex with men (MSM) are also exposed through sexual behavior.

**Objective:** This research aims to estimate the HAV seroprevalence and its predictors among young MSM.

**Methods:** This cross-sectional study, conducted in Rio de Janeiro/Brazil, included participants sequentially between 2019 and 2022. Inclusion criteria were cisgender males aged 18 to 35 who reported sex with men in the previous six months, had never received the HAV vaccine, and had no chronic immune-mediated conditions. Predictors represented sexual practices, use of different substances, sexually transmitted infections, signs or symptoms, water consumption and sanitation, and socio-demographic. The outcome of all the analyses was HAV serology (positive vs. negative). The main analysis compared the prediction performances of the random forest regression model with penalized GLM with internal cross-validation or bootstrap.

**Results:** The HAV seroprevalence was 39.6%. The compared models included from 16 to 42 predictors, always representing all predictor dimensions. “Chemsex” was identified as a predictor of HAV seroprevalence. A generation effect, interactions among race, education, and source of income were also identified as HAV predictors. The best model had excellent calibration and a moderate discrimination with an area under the ROC curve of 0.714 and an R^2^ of 0.187. We provide a web calculator at https://pedrobrasil.shinyapps.io/INDWELL/.

**Conclusion:** This study reveals a high susceptibility of cisgender young MSM to HAV infection. These findings highlight the enduring impact of socioeconomic inequities on enteric virus exposure and underscore the need for targeted public health strategies. One may use the prediction model to estimate the risk of seropositivity/susceptibility to HAV in aid of deciding either to perform a HAV test or to vaccinate against HAV, or even to estimate HAV seroprevalence in MSM populations where the HAV test is not easily available.

**HIGHLIGHTS:**

- The susceptibility to Hepatitis A infection among men who have sex with men (MSM) in Rio de Janeiro is approximately 60%.
- Usual exposures to Hepatitis A, such as socio-economic, sanitary, and the generation effect, are linked to Hepatitis A seroprevalence similarly in the MSM population.
- In addition to the traditional exposures to Hepatitis A virus, there is evidence linking directly sexual behaviors and preferences, and use of drugs to engaging in sex (“chemsex”) to Hepatitis A seropositivity.
- We developed an instrument to predict Hepatitis A seropositivity among men who have sex with men with good performance.

**ETHICS STATEMENT:** All procedures were performed in compliance with the good clinical practice document of the Americas (PAHO) and the Brazilian national Resolution n° 510/2016 and Resolution n° 466/2012 from the National Health Council (Conselho Nacional de Saúde – CNS). The institutional Ethics Committee (Instituto Nacional de Infectologia Evandro Chagas - Fundação Oswaldo Cruz) approved this research on March 22^nd^, 2018, with the number CAAE 80901917.0.0000.5262 (accessible at https://plataformabrasil.saude.gov.br/login.jsf). All participants gave written informed consent, and the privacy rights of human subjects were always observed.

## INTRODUCTION

Hepatitis A is an acute viral infection primarily transmitted via the fecal–oral route or through the ingestion of contaminated food or water. The incubation period ranges from 15 to 50 days, with a mean of approximately one month, and is inversely correlated with the infectious dose.[1] Clinical presentations vary from asymptomatic infection to severe illness. While older children and adults are more likely to manifest symptoms, particularly jaundice, younger children are often asymptomatic, with jaundice being uncommon.[2] Atypical forms (including cholestatic hepatitis and extrahepatic manifestations) may arise, especially in cases of prolonged illness. Fulminant hepatitis, the most severe form, occurs predominantly in older individuals and those with pre-existing liver disease, with a case fatality rate nearing 70% despite advances in intensive care.[3]

The hepatitis A virus (HAV) is highly resilient in the environment, facilitating extended transmission in areas with substandard sanitation.[4] Historically, this has led to high endemicity in low-income countries. However, urbanization and improvements in sanitation, hygiene, and socioeconomic conditions have driven a transition toward lower endemicity in many regions of the Global South, even where universal vaccination is not yet widespread.[5–7]

The World Health Organization (WHO) classifies hepatitis A endemicity based on age-specific seroprevalence of anti-HAV IgG antibodies, which indicate lifelong immunity. In high-endemicity regions, ≥90% of individuals acquire immunity by age 10 through early exposure.[2] Conversely, recent epidemiological studies further confirm that as regions transition from high to intermediate endemicity, the proportion of children with anti-HAV IgG by age 10 falls below this threshold, and the age at which 90% seroprevalence is reached shifts to adolescence or adulthood.[8–12] Additionally, in low-endemicity settings, infections are more frequent among specific groups, such as travelers, injection drug users, and men who have sex with men (MSM), reflecting a larger proportion of susceptible adults.[13–17]

Brazil has similarly undergone substantial epidemiological changes, with regional heterogeneity in transmission patterns driven by sanitation and urban development.[4] Between 2000 and 2022, children under 10 years of age accounted for 52.4% of all hepatitis A cases. Remarkably, from 2014 to 2022, the incidence among children aged 5 to 9 years declined by 99.2%. Nonetheless, since 2016, incidence rates have plateaued after three decades of sustained decline. In wealthier municipalities, outbreaks among young adults now represent a growing public health concern. Subgenotype IA remains predominant; however, recent molecular studies reveal the importation of strains during mass events, with 85.7% of circulating strains between 2017 and 2019 linked to MSM-associated outbreaks in Europe and Asia.[5,18]

Temporal patterns in Brazil exhibit two divergent trends: a continued decline in the North, Northeast, and Central-West regions, especially post-2014, and a stabilization or increase in the Southeast and South, marked by periodic outbreaks predominantly affecting adults aged 19 to 40 years.[18]

The incorporation of hepatitis A vaccination into Brazil’s national immunization program in 2014, offered as a single dose at 15 months of age, was a critical milestone. However, this first vaccinated cohort has not yet reached adulthood. Consequently, a significant portion of the adult population remains susceptible, particularly in urban areas such as Rio de Janeiro, where reasonable sanitation infrastructure is heterogeneous. The immunity landscape and associated risks among MSM in urban Brazil remain insufficiently characterized. Given the pervasive stigma and discrimination faced by LGBTQIA+ populations, this study aims to estimate the HAV seroprevalence and its predictors among young MSM. Such insights may help dispel misconceptions around sexual health practices and inform targeted prevention efforts for a population not yet reached by vaccine-induced protection.

## METHODS

### Design and settings

This cross-sectional study was conducted at the Reference Center for Special Immunobiologicals (CRIE), affiliated with Instituto Nacional de Infectologia Evandro Chagas / Fundação Oswaldo Cruz (INI-FIOCRUZ). INI-FIOCRUZ is an institute dedicated to research, teaching, and healthcare for infectious diseases, featuring a tertiary hospital with over 120 beds and outpatient clinics. Patients attending INI-FIOCRUZ usually live in the Rio de Janeiro metropolitan area and neighboring cities.

### Participants

Recruitment initially targeted venues such as gay nightclubs and events, but it was later adjusted. Participants were ultimately recruited at the center while seeking travel consultations, PrEP, or other services. Recruitment was performed sequentially between December 2019 and February 2022.

Eligible participants were cisgender males aged 18 to 35 who reported sex with men in the previous six months, had never received the HAV vaccine, and had no chronic immune-mediated conditions.

### Predictors

The predictors were verified regarding 6 different dimensions, including (1) sexual practices in the previous 5 years, sexual habits in different places, (2) use of different licit or illicit substances, (3) diagnosis of sexually transmitted infections in the previous 12 months, (4) signs or symptoms possibly related to HAV infection, history of traveling and travel exposures to HAV, (5) exposures related to water consumption and sanitation, and (6) socio-demographic characteristics such race and education..

Some definitions of some potential predictors: (1) Chemsex: intentional combination of psychoactive substances for the purpose of initiating, prolonging, or enhancing the sexual experience; (2) Cruising: active pursuit of random sexual partners in public or collective settings; (3) cruising bar: indoor establishment intended for cruising; (4) dark room: indoor environments with little or no lighting where anonymous and blind sex is practiced; (5) ecstasy: methylenedioxymethamphetamine, a synthetic psychoactive drug of the amphetamine family, used to obtain a tonic and euphoric effect; (6) fingering: sexual activity involving the insertion of fingers or toes into the partner’s anal opening; (7) fisting: sexual activity involving the insertion of hands beyond the fist, possibly reaching the more proximal segments of the upper limbs, into the partner’s anal opening; (8) Gi: Gamma-hydroxybutyrate, a natural neurotransmitter used as a synthetic psychoactive drug to obtain an aphrodisiac or sedative effect; (9) Pool party: a social event with a recreational purpose that takes place in a communal setting with a swimming pool, which may or may not be used by participants; (10) poppers: a generic name for volatile substances from the alkylnitrite chemical group, used for drug use and as sexual facilitators by promoting anal sphincter relaxation; (11) rave: a type of party usually lasting more than 12 hours, usually held away from urban centers and characterized by a predominance of electronic music; (12) scat: a sexual practice involving the intentional observation, handling, or ingestion of feces for pleasure.

### Outcome

The outcome of all the analyses was HAV serology (positive vs. negative). Participants provided a 10 mL blood sample. A 50 µL aliquot was tested immediately using the CTK Biotech Onsite™ LFIA to detect IgM and IgG anti-HAV antibodies. Indeterminate or positive LFIA-IgM results triggered confirmatory testing with CLIA. Participants negative for both IgM and IgG received the HAV vaccine unless contraindicated. Those ineligible for vaccination were counseled and exited the study. Serum samples were stored at −20°C and later tested by CLIA (DiaSorin LIAISON™) for total anti-HAV antibodies. Discordant results were reported, and participants were followed accordingly. Validation studies show LFIA achieves 96.1% concordance with the WHO-standard ELISA for IgM (specificity: 96.7%; sensitivity: 94.8%) and 98.0% for IgG (specificity: 94.7%; sensitivity: 100%) [14]. VAQTA (inactivated hepatitis A adsorbed vaccine), donated by Merck Sharp & Dohme LLC, was offered to seronegative participants.

### Analysis Plan

Data were stored in RedCap without identifiers and analyzed using R-project with packages Hmisc, mice, epitools, glmnet, glmnetUtils. randomForest, rfUtilities, givitiR, and Uncertaininterval. Missing data were explored and imputed using multiple imputation chained equations. Missing data assessments are available as supplemental. (Figure S1 to Figure S9) Categorical variables were recoded and, if necessary, collapsed. Continuous variables were assessed for distribution and outliers.

Descriptive statistics were computed. As part of the initial approach, prevalence ratios of HAV seropositivity were estimated within variable classes. For continuous variables, prevalence ratios were estimated using generalized linear models (GLM) with a Poisson distribution and exponentiation of the regression coefficient. For the continuous variables, the interpretation of the prevalence ratio should be an increment in prevalence for each unit of increment of the variable.

All the continuous predictors were tested as such (with no categorization such as normal vs. abnormal value limits) or with functional form transformations. The full model initially tested was year of birth, age, race/color (self-declared), place of birth, living place, marital status, education level, main source of income,” non-mineral, bottled, or treated water”, receives piped water, sewerage system in network, travel to another state, travel abroad, household contact with individual with acute hepatitis, leisure contact with individual with acute hepatitis, lives with child(ren) up to 12 years old, number of individuals in the same household, “lodging, fraternity/sorority house, barracks, or asylum”, sexually transmitted infections in the previous 12 months, number of sexual partners in the last 6 months, number of people kissed on the mouth in the last 6 months, receptive anal sex (passive), oral-genital sex with penis, oral-genital sex with mouth, oral-anal sex with mouth, scatological, insertive fisting, anal use of sex toys, group sex, receiving money for sex, paying for sex, anonymous sex or during cruising, sex parties or events, sex under the influence of drugs, suffering sexual abuse, “sauna, bathhouse, or cruising bar”, “nightclub, club, or other leisure places”, open public places, public restrooms, tobacco, alcohol, marijuana, cocaine, lysergic acid, LSD, acid, “ecstasy, pills, and similar”, “crystal, MD, michael douglas or similar”, ketamine, “Gi, GHB”, poppers, “solvents, inc. perfume inhalant”, psychotropics without medical prescription, anabolic steroids, other illicit substances, “HIV PrEP, Truvada”, vomiting, fecal acholia, “Fever + symptoms”. Some polynomials and interactions were tested.

The GLM regression approach was conducted with a cross-validated, elastic, and generalized linear model with a Poisson distribution. This approach enables the simultaneous penalization of the model, avoiding overfitting and optimism, using a lasso-type selection and ridge-type penalization.[19,20] Therefore, the predictors with no contribution to the predictions had their coefficients shrunk to 0. This approach does not drop the baseline level for factors, so that the regularization process shrinks the fitted coefficients toward the overall mean rather than toward the estimate for the baseline predictor level. The final model was extracted from 5 cross-validated folds, the fold with the least mean squared error. The final model coefficients were converted into prevalence ratios. Additionally, a random forest regression was performed with the following steps: tuning the number of variables randomly sampled as candidates at each split, adjusting the model with 2000 trees, and performing bias correction, selecting variables with Murphy model selection approach (based on variable importance),[21,22] estimate performance measurements including permutation cross-validation with 2000 bootstrap resampling.

Model performance measurement included calibration and discrimination, such as area under the ROC (receiver operating characteristics) curve (AUC - area under the curve), R^2^, Brier score, model intercept, model slope, and errors (distances) from the predicted probabilities to observed probabilities. Decision limits were estimated using the “uncertain interval” method,[23] which allows for an intermediate uncertain range of probabilities. The model was implemented on an online calculator using the shiny and shinydashboardPlus packages, allowing users to estimate individual probabilities at https://pedrobrasil.shinyapps.io/INDWELL/.

### Sample size

The male population in the municipality of Rio de Janeiro was about 2.96 million according to the 2010 census. It is estimated that 20% of the city’s male population identifies as gay or bisexual. Therefore, the estimated target population was 590,000 individuals. Assuming a 40% seropositivity rate, a 5% margin of error, and a 95% confidence level, we calculated a sample size of approximately 300 individuals to estimate HAV seroprevalence.

## RESULTS

Three hundred and three cisgender MSM participants aged between 18 and 35 were enrolled in the study. The HAV seroprevalence was estimated at 39.6% [95% CL 34.3%-45.2%]. The mean age of the participants was 27.83 years (SD = 4.37), with a median of 28 years and a range of 18 to 35 years. The age distribution was approximately symmetric and very similar among seropositive and seronegative subjects. Year of birth wasn’t a significant factor, but visually, as expected, it showed a slight decrease in point prevalence ratio. One can see this trend most clearly between 1991 and 2000. The mean for cohabitants was about 2.16, indicating most participants lived with others. Most participants reported having access to basic sanitation (e.g., piped water, sewage system), but there was variability across municipalities. Seropositivity was higher among Black and mixed-race individuals. Other potential predictors with higher HAV crude seroprevalence included widowhood, elementary education, being born in the Northeast, and household contact with an individual with acute hepatitis or during leisure activities. (Table 1).

**Table 1:** Socio-demographic and potential exposure characteristics by Hepatitis A serological status.

|  | Negative<br>(N=184) | Positive<br>(N=119) | Overall<br>(N=303) | Prevalence<br>Ratio |
| --- | --- | --- | --- | --- |
| **Year of birth** |  |  |  |  |
| Mean (SD) | 1992.92 (4.04) | 1992.16 (3.64) | 1992.62 (3.90) | 0.991 [0.967-1.016] |
| Median [IQR] | 1993.00<br>[1989.00,1996.00] | 1992.00<br>[1988.50,1995.00] | 1992.00<br>[1989.00,1996.00] |  |
| **Year of birth** |  |  |  |  |
| 1988 | 45 (24.5%) | 30 (25.2%) | 75 (24.8%) | 1.000 [NA-NA] |
| 1989 | 13 (7.1%) | 9 (7.6%) | 22 (7.3%) | 1.023 [0.576-1.815] |
| 1990 | 10 (5.4%) | 8 (6.7%) | 18 (5.9%) | 1.111 [0.618-1.997] |
| 1991 | 5 (2.7%) | 10 (8.4%) | 15 (5.0%) | 1.667 [1.060-2.621] |
| 1992 | 13 (7.1%) | 11 (9.2%) | 24 (7.9%) | 1.146 [0.684-1.919] |
| 1993 | 13 (7.1%) | 8 (6.7%) | 21 (6.9%) | 0.952 [0.517-1.756] |
| 1994 | 15 (8.2%) | 8 (6.7%) | 23 (7.6%) | 0.870 [0.466-1.624] |
| 1995 | 15 (8.2%) | 11 (9.2%) | 26 (8.6%) | 1.058 [0.624-1.793] |
| 1996 | 12 (6.5%) | 7 (5.9%) | 19 (6.3%) | 0.921 [0.480-1.766] |
| 1997 | 14 (7.6%) | 7 (5.9%) | 21 (6.9%) | 0.833 [0.428-1.621] |
| 1998 | 11 (6.0%) | 4 (3.4%) | 15 (5.0%) | 0.667 [0.275-1.613] |
| 1999 | 8 (4.3%) | 3 (2.5%) | 11 (3.6%) | 0.682 [0.250-1.861] |
| 2000 | 6 (3.3%) | 1 (0.8%) | 7 (2.3%) | 0.357 [0.057-2.239] |
| 2001 | 3 (1.6%) | 1 (0.8%) | 4 (1.3%) | 0.625 [0.112-3.490] |
| 2002 | 1 (0.5%) | 1 (0.8%) | 2 (0.7%) | 1.250 [0.304-5.137] |
| **Age** |  |  |  |  |
| Mean (SD) | 27.51 (4.42) | 28.39 (4.39) | 27.85 (4.42) | 1.008 [0.986-1.030] |
| Median [IQR] | 27.00 [24.00,31.00] | 29.00 [25.00,32.00] | 28.00 [24.00,31.00] |  |
| **Age** |  |  |  |  |
| [18,25] | 63 (34.2%) | 32 (26.9%) | 95 (31.4%) | 1.000 [NA-NA] |
| (25,30] | 62 (33.7%) | 49 (41.2%) | 111 (36.6%) | 1.311 [0.922-1.862] |
| (30,35] | 59 (32.1%) | 37 (31.1%) | 96 (31.7%) | 1.144 [0.783-1.671] |
| Missing | 0 (0%) | 1 (0.8%) | 1 (0.3%) |  |
| **Race/Color (Self-declared)** |  |  |  |  |
| White | 88 (47.8%) | 27 (22.7%) | 115 (38.0%) | 1.000 [NA-NA] |
| Black | 37 (20.1%) | 50 (42.0%) | 87 (28.7%) | 2.448 [1.680-3.566] |
| Yellow | 2 (1.1%) | 2 (1.7%) | 4 (1.3%) | 2.130 [0.757-5.989] |
| Indigenous | 1 (0.5%) | 0 (0%) | 1 (0.3%) | 0.000 [0.000-NaN] |
| Mixed-race | 56 (30.4%) | 40 (33.6%) | 96 (31.7%) | 1.775 [1.182-2.664] |
| **Marital status** |  |  |  |  |
| Single | 159 (86.4%) | 103 (86.6%) | 262 (86.5%) | 1.000 [NA-NA] |
| Married | 24 (13.0%) | 15 (12.6%) | 39 (12.9%) | 0.978 [0.640-1.496] |
| Widowed | 0 (0%) | 1 (0.8%) | 1 (0.3%) | 2.544 [2.188-2.957] |
| Separated | 1 (0.5%) | 0 (0%) | 1 (0.3%) | 0.000 [0.000-NaN] |
| **Education level** |  |  |  |  |
| Complete high | 59 (32.1%) | 43 (36.1%) | 102 (33.7%) | 1.000 [NA-NA] |
| Incomplete elementary | 2 (1.1%) | 4 (3.4%) | 6 (2.0%) | 1.581 [0.859-2.910] |
| Complete elementary | 0 (0%) | 3 (2.5%) | 3 (1.0%) | 2.372 [1.890-2.978] |
| Incomplete high | 3 (1.6%) | 5 (4.2%) | 8 (2.6%) | 1.483 [0.828-2.656] |
| Incomplete college | 53 (28.8%) | 26 (21.8%) | 79 (26.1%) | 0.781 [0.529-1.151] |
| Complete college | 47 (25.5%) | 32 (26.9%) | 79 (26.1%) | 0.961 [0.677-1.365] |
| Incomplete postgraduate | 9 (4.9%) | 1 (0.8%) | 10 (3.3%) | 0.237 [0.036-1.544] |
| Complete postgraduate | 11 (6.0%) | 5 (4.2%) | 16 (5.3%) | 0.741 [0.346-1.587] |
| **Main source of income** |  |  |  |  |
| Employed | 99 (53.8%) | 76 (63.9%) | 175 (57.8%) | 1.000 [NA- |
|  |  |  |  | NA] |
| Student | 38 (20.7%) | 4 (3.4%) | 42 (13.9%) | 0.219 [0.085-0.566] |
| Self-employed | 29 (15.8%) | 22 (18.5%) | 51 (16.8%) | 0.993 [0.695-1.420] |
| No source of income | 15 (8.2%) | 10 (8.4%) | 25 (8.3%) | 0.921 [0.554-1.532] |
| Other | 3 (1.6%) | 7 (5.9%) | 10 (3.3%) | 1.612 [1.039-2.502] |
| **Place of birth** |  |  |  |  |
| Southeast | 164 (89.1%) | 85 (71.4%) | 249 (82.2%) | 1.000 [NA-NA] |
| Northeast | 13 (7.1%) | 32 (26.9%) | 45 (14.9%) | 2.083 [1.616-2.685] |
| North | 3 (1.6%) | 2 (1.7%) | 5 (1.7%) | 1.172 [0.395-3.476] |
| Midwest | 1 (0.5%) | 0 (0%) | 1 (0.3%) | 0.000 [0.000-NaN] |
| South | 3 (1.6%) | 0 (0%) | 3 (1.0%) | 0.000 [0.000-NaN] |
| **Municipality of residence** |  |  |  |  |
| Rio de Janeiro city | 139 (75.5%) | 101 (84.9%) | 240 (79.2%) | 1.000 [NA-NA] |
| Metropolitan area | 45 (24.5%) | 18 (15.1%) | 63 (20.8%) | 0.679 [0.447-1.031] |
| **Non-mineral, non-bottled, or non-treated water?** |  |  |  |  |
| No | 79 (42.9%) | 45 (37.8%) | 124 (40.9%) | 1.000 [NA-NA] |
| Yes | 96 (52.2%) | 69 (58.0%) | 165 (54.5%) | 1.152 [0.858-1.547] |
| Missing | 9 (4.9%) | 5 (4.2%) | 14 (4.6%) |  |
| **Receives piped water?** |  |  |  |  |
| No | 7 (3.8%) | 7 (5.9%) | 14 (4.6%) | 1.000 [NA-NA] |
| Yes | 175 (95.1%) | 112 (94.1%) | 287 (94.7%) | 0.780 [0.453-1.344] |
| Missing | 2 (1.1%) | 0 (0%) | 2 (0.7%) |  |
| **Sewerage system in network?** |  |  |  |  |
| No | 11 (6.0%) | 8 (6.7%) | 19 (6.3%) | 1.000 [NA-NA] |
| Yes | 166 (90.2%) | 105 (88.2%) | 271 (89.4%) | 0.920 [0.532- |
|  |  |  |  | 1.592] |
| Missing | 7 (3.8%) | 6 (5.0%) | 13 (4.3%) |  |
| **Travel to another state?** |  |  |  |  |
| No | 93 (50.5%) | 71 (59.7%) | 164 (54.1%) | 1.000 [NA-NA] |
| Yes | 91 (49.5%) | 48 (40.3%) | 139 (45.9%) | 0.798 [0.598-1.064] |
| **Abroad travel?** |  |  |  |  |
| No | 170 (92.4%) | 116 (97.5%) | 286 (94.4%) | 1.000 [NA-NA] |
| Yes | 14 (7.6%) | 3 (2.5%) | 17 (5.6%) | 0.435 [0.154-1.227] |
| **Household contact with individual with acute hepatitis?** |  |  |  |  |
| No | 132 (71.7%) | 74 (62.2%) | 206 (68.0%) | 1.000 [NA-NA] |
| Yes | 0 (0%) | 2 (1.7%) | 2 (0.7%) | 2.784 [2.320-3.341] |
| Missing | 52 (28.3%) | 43 (36.1%) | 95 (31.4%) |  |
| **Leisure contact with individual with acute hepatitis?** |  |  |  |  |
| No | 99 (53.8%) | 70 (58.8%) | 169 (55.8%) | 1.000 [NA-NA] |
| Yes | 0 (0%) | 1 (0.8%) | 1 (0.3%) | 2.414 [2.018-2.888] |
| Missing | 85 (46.2%) | 48 (40.3%) | 133 (43.9%) |  |
| **Lives with child(ren) up to 12 years old?** |  |  |  |  |
| No | 154 (83.7%) | 91 (76.5%) | 245 (80.9%) | 1.000 [NA-NA] |
| Yes | 26 (14.1%) | 25 (21.0%) | 51 (16.8%) | 1.320 [0.955-1.824] |
| Missing | 4 (2.2%) | 3 (2.5%) | 7 (2.3%) |  |
| **Number of individuals in the same household** |  |  |  |  |
| Mean (SD) | 2.14 (1.90) | 2.19 (1.81) | 2.16 (1.86) | 1.003 [0.951-1.054] |
| Median [IQR] | 2.00 [1.00,3.00] | 2.00 [1.00,3.00] | 2.00 [1.00,3.00] |  |
| **Number of individuals in the same household** |  |  |  |  |
| 0 | 31 (16.8%) | 16 (13.4%) | 47 (15.5%) | 1.000 [NA-NA] |
| 1 | 49 (26.6%) | 38 (31.9%) | 87 (28.7%) | 1.283 [0.807- |
|  |  |  |  | 2.041] |
| 2 | 40 (21.7%) | 20 (16.8%) | 60 (19.8%) | 0.979 [0.573-1.672] |
| 3 | 28 (15.2%) | 22 (18.5%) | 50 (16.5%) | 1.292 [0.779-2.144] |
| 4 | 22 (12.0%) | 10 (8.4%) | 32 (10.6%) | 0.918 [0.479-1.758] |
| ≥5 | 14 (7.6%) | 13 (10.9%) | 27 (8.9%) | 1.414 [0.809-2.472] |
| **Lodging,**<br>**fraternity/sorority**<br>**house, barracks, or**<br>**asylum?** |  |  |  |  |
| No | 175 (95.1%) | 115 (96.6%) | 290 (95.7%) | 1.000 [NA-NA] |
| Yes | 7 (3.8%) | 3 (2.5%) | 10 (3.3%) | 0.757 [0.290-1.971] |
| Missing | 2 (1.1%) | 1 (0.8%) | 3 (1.0%) |  |

IQR = interquartile range; SD = Standard deviation; Prevalence ratio for continuous variables is estimated through GLM with Poisson distribution with robust 95% confidence intervals. For categorical variables 1 is the reference category. NA = not assigned.

Regarding sexual behavior, the number of reported sexual partners exhibited substantial variability. The number of sexual partners in the previous 6 months was considered very high, ranging from 1 to 200 partners, with a mean of 10.39. The median number of partners was 4, suggesting a skewed distribution with a minority of participants reporting very high partner counts. The use of PrEP was common. For sexual behavior, initial analysis identified oral-genital contact, use of sex toys anally, group sex, and practicing sex in public restrooms as potential predictors with significantly increased HAV seroprevalence. (Table 2)

**Table 2:** Sexual preferences and behaviors by Hepatitis A serological status.

|  | Negative<br>(N=184) | Positive<br>(N=119) | Overall<br>(N=303) | Prevalence<br>Ratio |
| --- | --- | --- | --- | --- |
| **Sexual contact with individual with acute hepatitis?** |  |  |  |  |
| No | 104 (56.5%) | 74 (62.2%) | 178 (58.7%) | 1.000 [NA-NA] |
| Yes | 1 (0.5%) | 1 (0.8%) | 2 (0.7%) | 1.203 [0.298-4.862] |
| Missing | 79 (42.9%) | 44 (37.0%) | 123 (40.6%) |  |
| **STI previous 12 months** |  |  |  |  |
| No | 129 (70.1%) | 78 (65.5%) | 207 (68.3%) | 1.000 [NA-NA] |
| Yes | 48 (26.1%) | 35 (29.4%) | 83 (27.4%) | 1.119 [0.823-1.521] |
| Missing | 7 (3.8%) | 6 (5.0%) | 13 (4.3%) |  |
| **Number of sexual partners in the last 6 months** |  |  |  |  |
| Mean (SD) | 11.27 (25.82) | 9.02 (13.64) | 10.38 (21.86) | 0.999 [0.994-1.003] |
| Median [IQR] | 4.00<br>[2.00,10.00] | 4.00<br>[1.00,10.00] | 4.00<br>[2.00,10.00] |  |
| **Number of people kissed on the mouth in the last 6 months** |  |  |  |  |
| Mean (SD) | 15.75 (35.42) | 11.75 (17.54) | 14.18 (29.74) | 0.999 [0.995-1.002] |
| Median [IQR] | 5.50<br>[2.00,15.00] | 5.00<br>[2.00,11.00] | 5.00<br>[2.00,15.00] |  |
| **Receptive anal sex (passive)** |  |  |  |  |
| Yes | 163 (88.6%) | 110 (92.4%) | 273 (90.1%) | 1.000 [NA-NA] |
| No | 21 (11.4%) | 9 (7.6%) | 30 (9.9%) | 0.745 [0.423-1.310] |
| **Oral-genital sex with penis** |  |  |  |  |
| Yes | 177 (96.2%) | 108 (90.8%) | 285 (94.1%) | 1.000 [NA-NA] |
| No | 7 (3.8%) | 11 (9.2%) | 18 (5.9%) | 1.613 [1.084-2.399] |
| **Oral-genital sex with mouth** |  |  |  |  |
| Yes | 176 (95.7%) | 104 (87.4%) | 280 (92.4%) | 1.000 [NA-NA] |
| No | 8 (4.3%) | 14 (11.8%) | 22 (7.3%) | 1.713 [1.206-2.433] |
| Missing | 0 (0%) | 1 (0.8%) | 1 (0.3%) |  |
| **Oral-anal sex with mouth** |  |  |  |  |
| Yes | 160 (87.0%) | 95 (79.8%) | 255 (84.2%) | 1.000 [NA-NA] |
| No | 23 (12.5%) | 24 (20.2%) | 47 (15.5%) | 1.371 [0.993-1.891] |
| Missing | 1 (0.5%) | 0 (0%) | 1 (0.3%) |  |
| **Scatological** |  |  |  |  |
| No | 179 (97.3%) | 115 (96.6%) | 294 (97.0%) | 1.000 [NA-NA] |
| Yes | 5 (2.7%) | 4 (3.4%) | 9 (3.0%) | 1.136 [0.540-2.392] |
| **Insertive fisting** |  |  |  |  |
| No | 161 (87.5%) | 105 (88.2%) | 266 (87.8%) | 1.000 [NA-NA] |
| Yes | 23 (12.5%) | 14 (11.8%) | 37 (12.2%) | 0.959 [0.618-1.487] |
| **Receptive fisting** |  |  |  |  |
| No | 167 (90.8%) | 116 (97.5%) | 283 (93.4%) | 1.000 [NA-NA] |
| Yes | 17 (9.2%) | 2 (1.7%) | 19 (6.3%) | 0.257 [0.069-0.960] |
| Missing | 0 (0%) | 1 (0.8%) | 1 (0.3%) |  |
| **Anal use of sex toys** |  |  |  |  |
| No | 111 (60.3%) | 87 (73.1%) | 198 (65.3%) | 1.000 [NA-NA] |
| Yes | 72 (39.1%) | 32 (26.9%) | 104 (34.3%) | 0.700 [0.504-0.973] |
| Missing | 1 (0.5%) | 0 (0%) | 1 (0.3%) |  |
| **Group sex** |  |  |  |  |
| No | 65 (35.3%) | 61 (51.3%) | 126 (41.6%) | 1.000 [NA-NA] |
| Yes | 119 (64.7%) | 57 (47.9%) | 176 (58.1%) | 0.669 [0.506-0.885] |
| Missing | 0 (0%) | 1 (0.8%) | 1 (0.3%) |  |
| **Receiving money for sex** |  |  |  |  |
| No | 165 (89.7%) | 106 (89.1%) | 271 (89.4%) | 1.000 [NA-NA] |
| Yes | 18 (9.8%) | 12 (10.1%) | 30 (9.9%) | 1.023 [0.644-1.624] |
| Missing | 1 (0.5%) | 1 (0.8%) | 2 (0.7%) |  |
| **Paying for sex** |  |  |  |  |
| No | 171 (92.9%) | 114 (95.8%) | 285 (94.1%) | 1.000 [NA-NA] |
| Yes | 13 (7.1%) | 5 (4.2%) | 18 (5.9%) | 0.694 [0.325-1.482] |
| **Anonymous sex or during cruising** |  |  |  |  |
| No | 114 (62.0%) | 72 (60.5%) | 186 (61.4%) | 1.000 [NA-NA] |
| Yes, always protected | 30 (16.3%) | 14 (11.8%) | 44 (14.5%) | 0.822 [0.514-1.314] |
| Yes | 36 (19.6%) | 28 (23.5%) | 64 (21.1%) | 1.130 [0.811-1.574] |
| Missing | 4 (2.2%) | 5 (4.2%) | 9 (3.0%) |  |
| **Sex parties or events** |  |  |  |  |
| No | 134 (72.8%) | 85 (71.4%) | 219 (72.3%) | 1.000 [NA-NA] |
| Yes | 50 (27.2%) | 33 (27.7%) | 83 (27.4%) | 1.024 [0.749- |
|  |  |  |  | 1.400] |
| Missing | 0 (0%) | 1 (0.8%) | 1 (0.3%) |  |
| **Sex under the influence of drugs** |  |  |  |  |
| No | 126 (68.5%) | 81 (68.1%) | 207 (68.3%) | 1.000 [NA-NA] |
| Yes | 58 (31.5%) | 37 (31.1%) | 95 (31.4%) | 0.995 [0.735-1.349] |
| Missing | 0 (0%) | 1 (0.8%) | 1 (0.3%) |  |
| **Suffering sexual abuse** |  |  |  |  |
| No | 173 (94.0%) | 111 (93.3%) | 284 (93.7%) | 1.000 [NA-NA] |
| Yes | 8 (4.3%) | 7 (5.9%) | 15 (5.0%) | 1.194 [0.682-2.091] |
| Missing | 3 (1.6%) | 1 (0.8%) | 4 (1.3%) |  |
| **Sauna, bathhouse, or cruising bar** |  |  |  |  |
| No | 109 (59.2%) | 77 (64.7%) | 186 (61.4%) | 1.000 [NA-NA] |
| Yes | 75 (40.8%) | 41 (34.5%) | 116 (38.3%) | 0.854 [0.633-1.152] |
| Missing | 0 (0%) | 1 (0.8%) | 1 (0.3%) |  |
| **Nightclub, club, or other leisure places** |  |  |  |  |
| No | 109 (59.2%) | 73 (61.3%) | 182 (60.1%) | 1.000 [NA-NA] |
| Yes | 74 (40.2%) | 44 (37.0%) | 118 (38.9%) | 0.930 [0.693-1.247] |
| Missing | 1 (0.5%) | 2 (1.7%) | 3 (1.0%) |  |
| **Open public places** |  |  |  |  |
| No | 96 (52.2%) | 73 (61.3%) | 169 (55.8%) | 1.000 [NA-NA] |
| Yes | 88 (47.8%) | 46 (38.7%) | 134 (44.2%) | 0.795 [0.594-1.063] |
| **Public restrooms** |  |  |  |  |
| No | 124 (67.4%) | 95 (79.8%) | 219 (72.3%) | 1.000 [NA-NA] |
| Yes | 60 (32.6%) | 24 (20.2%) | 84 (27.7%) | 0.659 [0.455-0.954] |
| **Public pool or pool party** |  |  |  |  |
| No | 169 (91.8%) | 110 (92.4%) | 279 (92.1%) | 1.000 [NA-NA] |
| Yes | 15 (8.2%) | 7 (5.9%) | 22 (7.3%) | 0.807 [0.430-1.513] |
| Missing | 0 (0%) | 2 (1.7%) | 2 (0.7%) |  |
| **Hand washing immediately after sex** |  |  |  |  |
| Yes | 128 (69.6%) | 85 (71.4%) | 213 (70.3%) | 1.000 [NA-NA] |
| No | 53 (28.8%) | 32 (26.9%) | 85 (28.1%) | 0.943 [0.685-1.298] |
| Missing | 3 (1.6%) | 2 (1.7%) | 5 (1.7%) |  |
| **HIV PrEP, Truvada** |  |  |  |  |
| No | 127 (69.0%) | 68 (57.1%) | 195 (64.4%) | 1.000 [NA-NA] |
| Yes | 57 (31.0%) | 46 (38.7%) | 103 (34.0%) | 1.281 [0.960-1.708] |
| Missing | 0 (0%) | 5 (4.2%) | 5 (1.7%) |  |

IQR = interquartile range; SD = Standard deviation; STI = sexually transmitted infection; PrEP = Pre-Exposure Prophylaxis; NA = not assigned; 1 is the reference category.

Both legal and illegal drug use were common. Prevalence varied notably by drug, ranging from 10% for psychotropics up to 87% for alcohol. Additionally, several drugs had a prevalence ratio pointing to a crude association with HAV seropositivity, including marijuana, LSD, ecstasy, and crystal. All the predictors decreased the HAV seroprevalence when compared to the absence of drug use. (Table 3). From the few signs and symptoms explored, jaundice was the least frequent; however, it was the only one to have an increased prevalence ratio to HAV seropositivity. (Table 4)

**Table 3:** Drug use and medication use by Hepatitis A serological status.

|  | Negative<br>(N=184) | Positive<br>(N=119) | Overall<br>(N=303) | Prevalence Ratio |
| --- | --- | --- | --- | --- |
| **Tobacco** |  |  |  |  |
| No | 113 (61.4%) | 81 (68.1%) | 194 (64.0%) | 1.000 [NA-NA] |
| Yes | 68 (37.0%) | 37 (31.1%) | 105 (34.7%) | 0.844 [0.620-1.148] |
| Missing | 3 (1.6%) | 1 (0.8%) | 4 (1.3%) |  |
| **Alcohol** |  |  |  |  |
| No | 25 (13.6%) | 13 (10.9%) | 38 (12.5%) | 1.000 [NA-NA] |
| Yes | 159 (86.4%) | 106 (89.1%) | 265 (87.5%) | 1.169 [0.734-1.861] |
| **Marijuana** |  |  |  |  |
| No | 73 (39.7%) | 72 (60.5%) | 145 (47.9%) | 1.000 [NA-NA] |
| Yes | 111 (60.3%) | 46 (38.7%) | 157 (51.8%) | 0.590 [0.440-0.791] |
| Missing | 0 (0%) | 1 (0.8%) | 1 (0.3%) |  |
| **Cocaine** |  |  |  |  |
| No | 151 (82.1%) | 97 (81.5%) | 248 (81.8%) | 1.000 [NA-NA] |
| Yes | 33 (17.9%) | 22 (18.5%) | 55 (18.2%) | 1.023 [0.714-1.464] |
| **Lysergic acid, LSD, acid** |  |  |  |  |
| No | 137 (74.5%) | 104 (87.4%) | 241 (79.5%) | 1.000 [NA-NA] |
| Yes | 45 (24.5%) | 15 (12.6%) | 60 (19.8%) | 0.579 [0.365-0.919] |
| Missing | 2 (1.1%) | 0 (0%) | 2 (0.7%) |  |
| **Ecstasy, pills, and similar** |  |  |  |  |
| No | 123 (66.8%) | 92 (77.3%) | 215 (71.0%) | 1.000 [NA-NA] |
| Yes | 61 (33.2%) | 26 (21.8%) | 87 (28.7%) | 0.698 [0.489-0.998] |
| Missing | 0 (0%) | 1 (0.8%) | 1 (0.3%) |  |
| **Crystal, MD, michael douglas or similar** |  |  |  |  |
| No | 137 (74.5%) | 107 (89.9%) | 244 (80.5%) | 1.000 [NA-NA] |
| Yes | 46 (25.0%) | 12 (10.1%) | 58 (19.1%) | 0.472 [0.280-0.796] |
| Missing | 1 (0.5%) | 0 (0%) | 1 (0.3%) |  |
| **Key, Ketamine** |  |  |  |  |
| No | 146 (79.3%) | 102 (85.7%) | 248 (81.8%) | 1.000 [NA-NA] |
| Yes | 38 (20.7%) | 17 (14.3%) | 55 (18.2%) | 0.752 [0.493-1.146] |
| **Gi, GHB** |  |  |  |  |
| No | 157 (85.3%) | 102 (85.7%) | 259 (85.5%) | 1.000 [NA-NA] |
| Yes | 27 (14.7%) | 17 (14.3%) | 44 (14.5%) | 0.981 [0.656-1.466] |
| **Crack and similar** |  |  |  |  |
| No | 181 (98.4%) | 119 (100%) | 300 (99.0%) | 1.000 [NA-NA] |
| Yes | 2 (1.1%) | 0 (0%) | 2 (0.7%) | 0.000 [0.000-NaN] |
| Missing | 1 (0.5%) | 0 (0%) | 1 (0.3%) |  |
| **Poppers** |  |  |  |  |
| No | 148 (80.4%) | 102 (85.7%) | 250 (82.5%) | 1.000 [NA-NA] |
| Yes | 36 (19.6%) | 17 (14.3%) | 53 (17.5%) | 0.786 [0.517-1.196] |
| **Solvents, inc. Perfume inhalant** |  |  |  |  |
| No | 147 (79.9%) | 106 (89.1%) | 253 (83.5%) | 1.000 [NA-NA] |
| Yes | 37 (20.1%) | 13 (10.9%) | 50 (16.5%) | 0.621 [0.380-1.013] |
| **Psychotropics without medical prescription** |  |  |  |  |
| No | 160 (87.0%) | 113 (95.0%) | 273 (90.1%) | 1.000 [NA-NA] |
| Yes | 23 (12.5%) | 6 (5.0%) | 29 (9.6%) | 0.500 [0.242-1.034] |
| Missing | 1 (0.5%) | 0 (0%) | 1 (0.3%) |  |
| **Anabolic steroids** |  |  |  |  |
| No | 159 (86.4%) | 100 (84.0%) | 259 (85.5%) | 1.000 [NA-NA] |
| Yes | 25 (13.6%) | 19 (16.0%) | 44 (14.5%) | 1.118 [0.771-1.623] |
| **Other illicit substances** |  |  |  |  |
| No | 169 (91.8%) | 114 (95.8%) | 283 (93.4%) | 1.000 [NA-NA] |
| Yes | 9 (4.9%) | 3 (2.5%) | 12 (4.0%) | 0.621 [0.231-1.671] |
| Missing | 6 (3.3%) | 2 (1.7%) | 8 (2.6%) |  |

IQR = interquartile range; SD = Standard deviation; 1 is the reference category. NA = not assigned; Gi,GHB = Gamma-hydroxybutyrate; Poppers = volatile substances from the alkyl nitrite chemical group; Crystal = methamphetamine; HIV = human immunodeficiency virus.

**Table 4:** Signs and symptoms by Hepatitis A serological status.

|  | Negative<br>(N=184) | Positive<br>(N=119) | Overall<br>(N=303) | Prevalence Ratio |
| --- | --- | --- | --- | --- |
| **Vomiting** |  |  |  |  |
| No | 160 (87.0%) | 104 (87.4%) | 264 (87.1%) | 1.000 [NA-NA] |
| Yes | 24 (13.0%) | 14 (11.8%) | 38 (12.5%) | 0.935 [0.601-1.456] |
| Missing | 0 (0%) | 1 (0.8%) | 1 (0.3%) |  |
| **Jaundice** |  |  |  |  |
| No | 181 (98.4%) | 113 (95.0%) | 294 (97.0%) | 1.000 [NA-NA] |
| Yes | 0 (0%) | 5 (4.2%) | 5 (1.7%) | 2.602 [2.251-3.007] |
| Missing | 3 (1.6%) | 1 (0.8%) | 4 (1.3%) |  |
| **Choluria** |  |  |  |  |
| No | 178 (96.7%) | 115 (96.6%) | 293 (96.7%) | 1.000 [NA-NA] |
| Yes | 6 (3.3%) | 2 (1.7%) | 8 (2.6%) | 0.637 [0.190-2.133] |
| Missing | 0 (0%) | 2 (1.7%) | 2 (0.7%) |  |
| **Fecal acholia** |  |  |  |  |
| No | 163 (88.6%) | 100 (84.0%) | 263 (86.8%) | 1.000 [NA-NA] |
| Yes | 17 (9.2%) | 14 (11.8%) | 31 (10.2%) | 1.188 [0.782-1.803] |
| Missing | 4 (2.2%) | 5 (4.2%) | 9 (3.0%) |  |
| **Fever + symptoms** |  |  |  |  |
| No | 167 (90.8%) | 113 (95.0%) | 280 (92.4%) | 1.000 [NA-NA] |
| Yes | 12 (6.5%) | 4 (3.4%) | 16 (5.3%) | 0.619 [0.262-1.465] |
| Missing | 5 (2.7%) | 2 (1.7%) | 7 (2.3%) |  |

1 is the reference category. NA = not assigned.

There were two main full models in the random forest approach and the GLM approach. The first without place of birth and living area, following the idea that, like this, the models would have a wider prediction applicability. Then, the same procedures of model adjustment were performed, including the place of birth and living area as initial predictors. The initial random forest model without the place of birth and the living area led to a final model with 31 predictors. Naming from the most important to the least important, they were: “Race/Color (Self-declared)”, “Main source of income”, “Marijuana”, “Group sex”, “Alcohol”, “Anal use of sex toys”, “Crystal, MD, michael douglas or similar”, “Education level”, “Number of individuals in the same household”, “HIV PrEP, Truvada”, “Lysergic acid, LSD, acid”, “Oral-genital sex with mouth”, “Receptive fisting”, “Receptive anal sex (passive)”, “Year of birth”, “Marital status”, “Solvents, inc. Perfume inhalant”, “Jaundice”, “Sex under the influence of drugs”, “Lives with child(ren) up to 12 years old?”, “Oral-anal sex with mouth”, “Abroad travel?”, “Sauna, bathhouse, or cruising bar”, “Travel to another state?”, “Poppers”, “Ecstasy, pills, and similar”, “Household contact with an individual with acute hepatitis?”, “Public pool or pool party”, “Choluria”, “Suffering sexual abuse”, “Crack and similar”. (Figure 1) One may see that predictors of all dimensions are represented (socio-demographic, sex behaviors and preferences, drug use, and signs and symptoms). The importance of the dropped predictors decreases steadily to negative values. (Figure S10)

**Figure 1:**
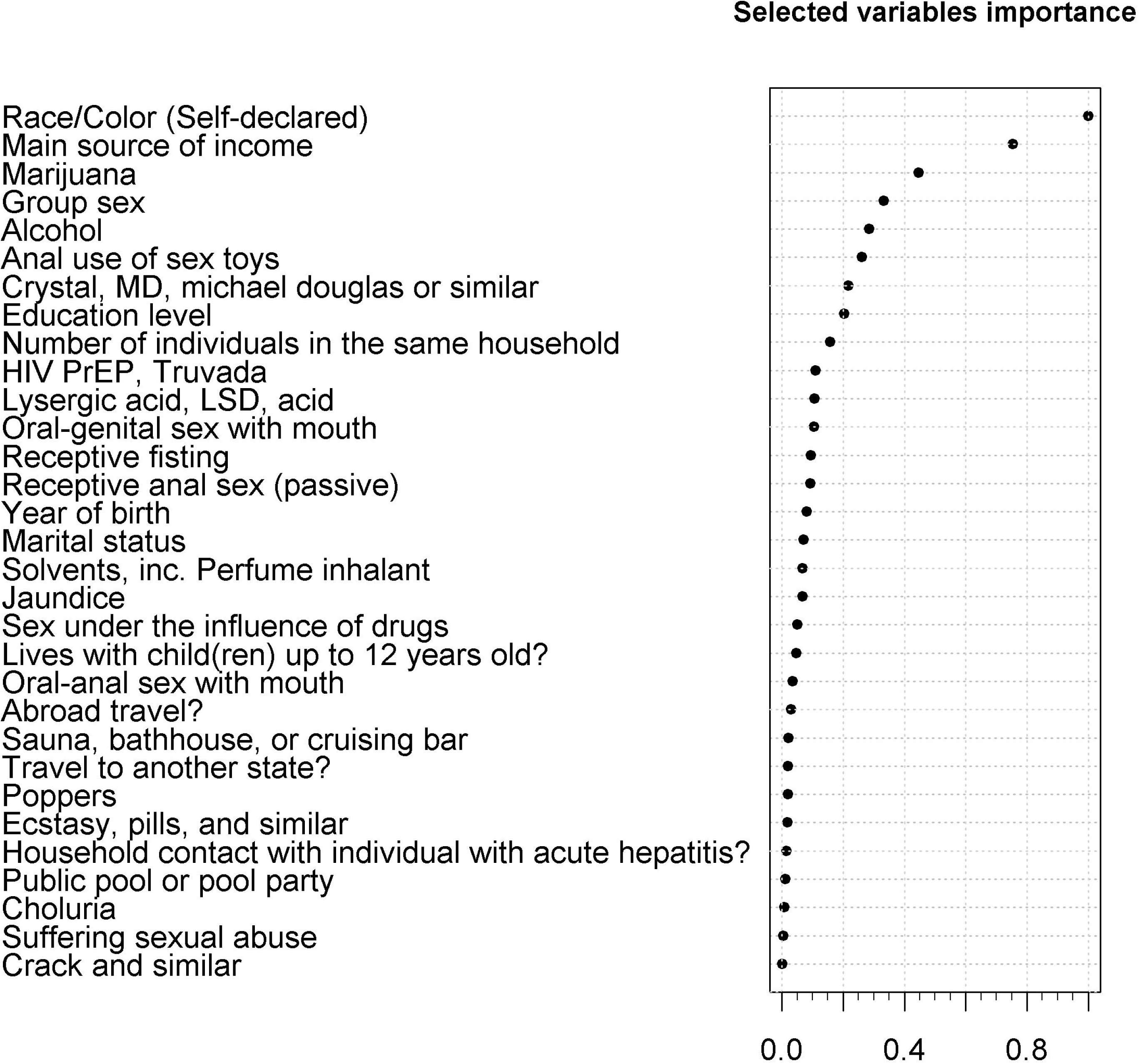

When compared to the former, the random forest model with the place of birth and the living place had fewer predictors (16), which are: “Place of birth”, “Race/Color (Self-declared)”, “Main source of income”, “Marijuana”, “Alcohol, “Crystal, MD, michael douglas or similar”, “Education level”, “Group sex”, “Anal use of sex toys”, “Lysergic acid, LSD, acid”, “Oral-genital sex with mouth”, “Sex under the influence of drugs”, “Marital status”, “Number of individuals in the same household”, “Jaundice”, “Receptive anal sex (passive)”. (Figure S11 and Figure S12) This approach found “place of birth” as the most important predictor and most of the important predictors of the former approach were also important in this second approach. Additionally, many of the less important predictors were dropped when place of birth was considered. Again, all predictor dimensions were represented.

The random forest model performance was considered moderate to good. The calibration was excellent, with an intercept and slope very close to 0 and 1, respectively, with grouped observations of 20 subjects showing little variation in the lower range of predictions, and a calibration belt without evidence of predictions significantly above or under the optimal diagonal, with reasonable average prediction error from the observed values. (Figures 2A and 2B) The final model demonstrated moderate discrimination, as indicated by a moderate area under the ROC curve and a good R². (Figure 2A) The bootstrap with cross-validation approach yielded overall good performance metrics, with moderate to good variance explained, a fit mean squared error, and near-zero cross-validation errors and variance. (Table 5) Although the ROC curve was slightly skewed to the left, the final model’s uncertain range of predictions was very narrow, between 0.43 and 0.46. (Figure 2C) The random forest approach with the place of birth had similar results; however, it was more accurate in discrimination, keeping excellent calibration, (Figure S13) including similar or slightly better cross-validation metrics.

**Figure 2:**
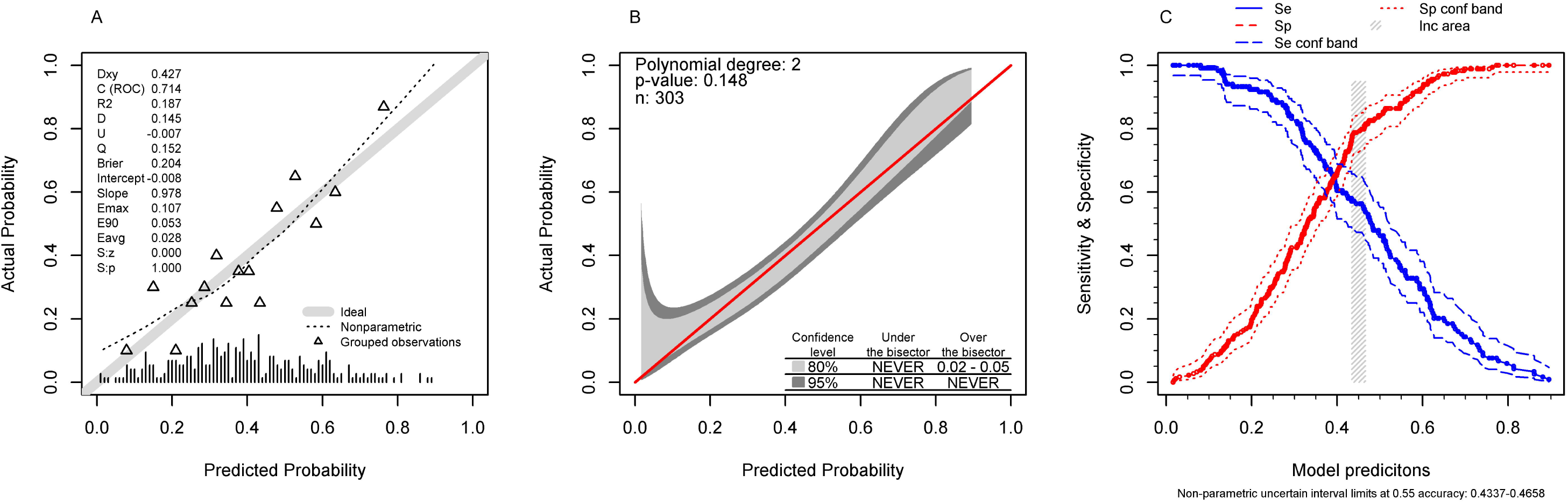

**Table 5:**
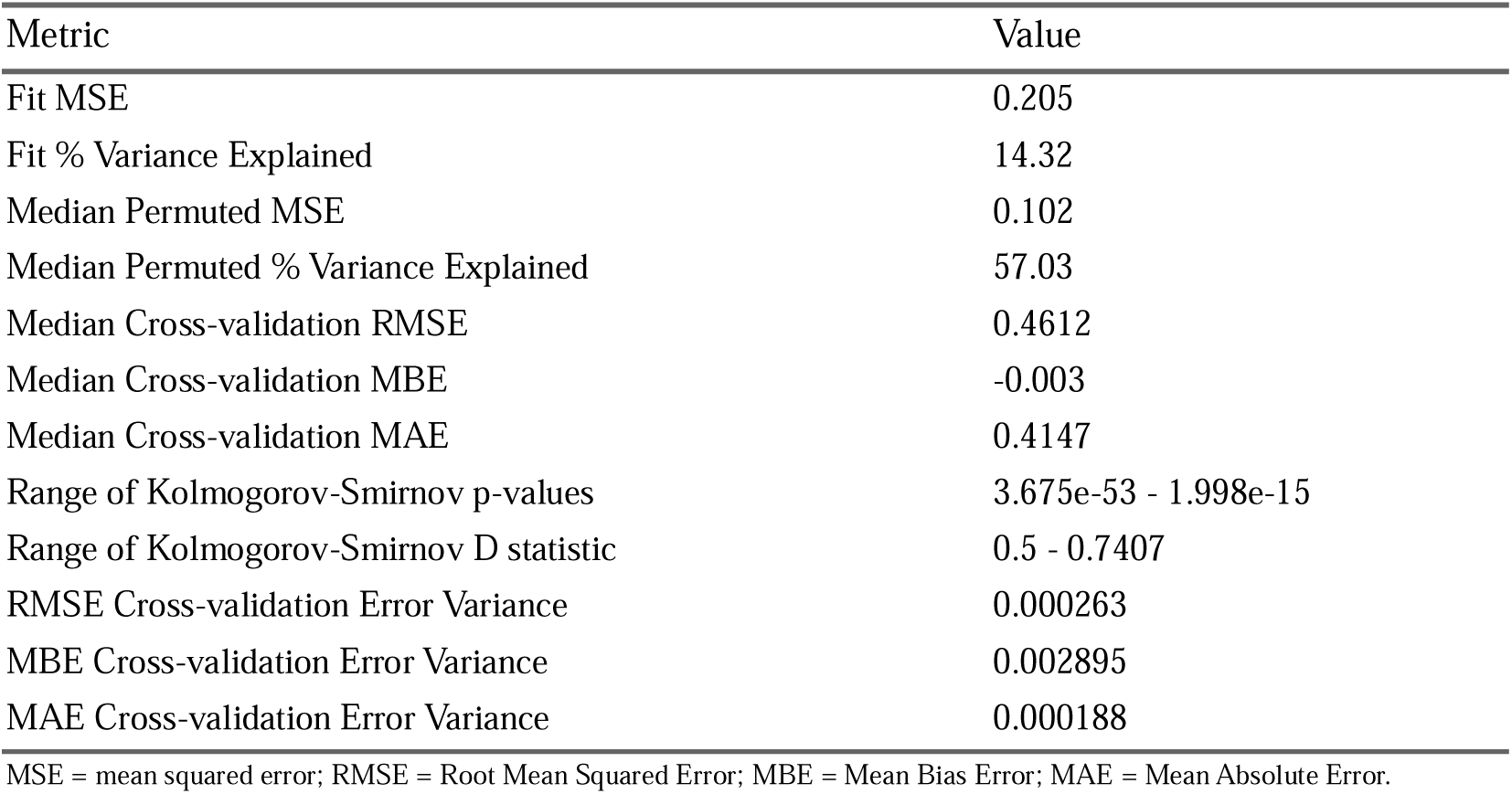
Random forest regression model cross-validation metrics to predict Hepatitis A sero-positivity.

The final GLM model without the place of birth had 41 predictors: “Age”, “Race/Color (Self-declared)”, “Marital status”, “Education level”, “Main source of income”, “Non-mineral, non-bottled, or non-treated water?”, “Receives piped water?”, “Travel to another state?”, “Abroad travel?”, “Household contact with individual with acute hepatitis?”, “Leisure contact with individual with acute hepatitis?”, “Lives with child(ren) up to 12 years old?”, “STI previous 12 months”, “Receptive anal sex (passive)”, “Oral-genital sex with penis”, “Oral-genital sex with mouth”, “Receptive fisting”, “Anal use of sex toys”, “Group sex”, “Paying for sex”, “Anonymous sex or during cruising”, “Sex parties or events”, “Sex under the influence of drugs”, “Suffering sexual abuse”, “Public restrooms”, “Alcohol”, “Marijuana”, “Cocaine”, “Lysergic acid, LSD, acid”, “Crystal, MD, michael douglas or similar”, “Crack and similar”, “Psychotropics without medical prescription”, “HIV PrEP, Truvada”, “Jaundice”, “Choluria”, “Fecal acholia”, “Fever + symptoms”. All predictor dimensions were represented in the final model. This result also identified that year of birth was significant as a polynomial transformation, meaning that year of birth did not have a linear relationship shape when predicting HAV sero-positivity. (Figure S14) In addition, this result also identified interactions of year of birth with age, despite the short period of recruitment, considering a possible generation effect. It also identified race interaction with education and race interaction with source of income. (Table S2)

The final GLM model with the place of birth and living area had 42 predictors, which are “Age”, “Race/Color (Self-declared)”, “Marital status”, “Education level”, “Main source of income”, “Place of birth”, “Municipality of residence”, “Non-mineral, non-bottled, or non-treated water?”, “Receives piped water?”, “Travel to another state?”, “Abroad travel?”, “Household contact with individual with acute hepatitis?”, “Leisure contact with individual with acute hepatitis?”, “Lives with child(ren) up to 12 years old?”, “STI previous 12 months”, “Receptive anal sex (passive)”, “Oral-genital sex with mouth”, “Receptive fisting”, “Anal use of sex toys”, “Group sex”, “Paying for sex”, “Anonymous sex or during cruising”, “Sex parties or events”, “Sex under the influence of drugs”, “Suffering sexual abuse”, “Public restrooms”, “Alcohol”, “Marijuana”, “Cocaine”, “Lysergic acid”, “LSD, acid”, “Crystal, MD, michael douglas or similar”, “Crack and similar”, “Psychotropics without medical prescription”, “HIV PrEP, Truvada”, “Jaundice”, “Choluria”, “Fecal acholia”, “Fever + symptoms”. Again, this approach included the year of birth non-linear relationship, interactions of year of birth with age, race with education, and race with source of income. The only predictor included in the previous GLM approach not retained here was “Oral-genital sex with penis”. (Table 6)

**Table 6:**
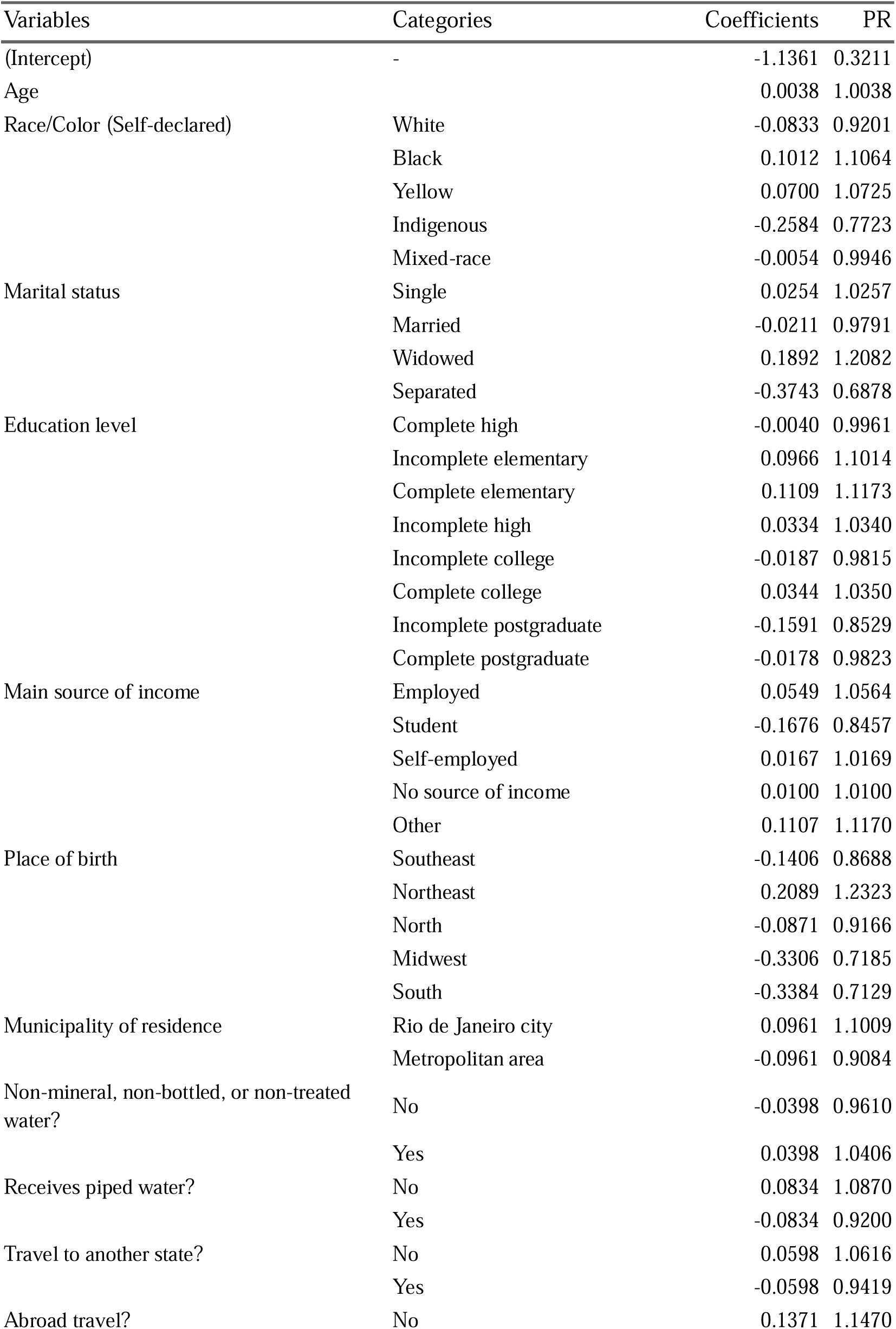
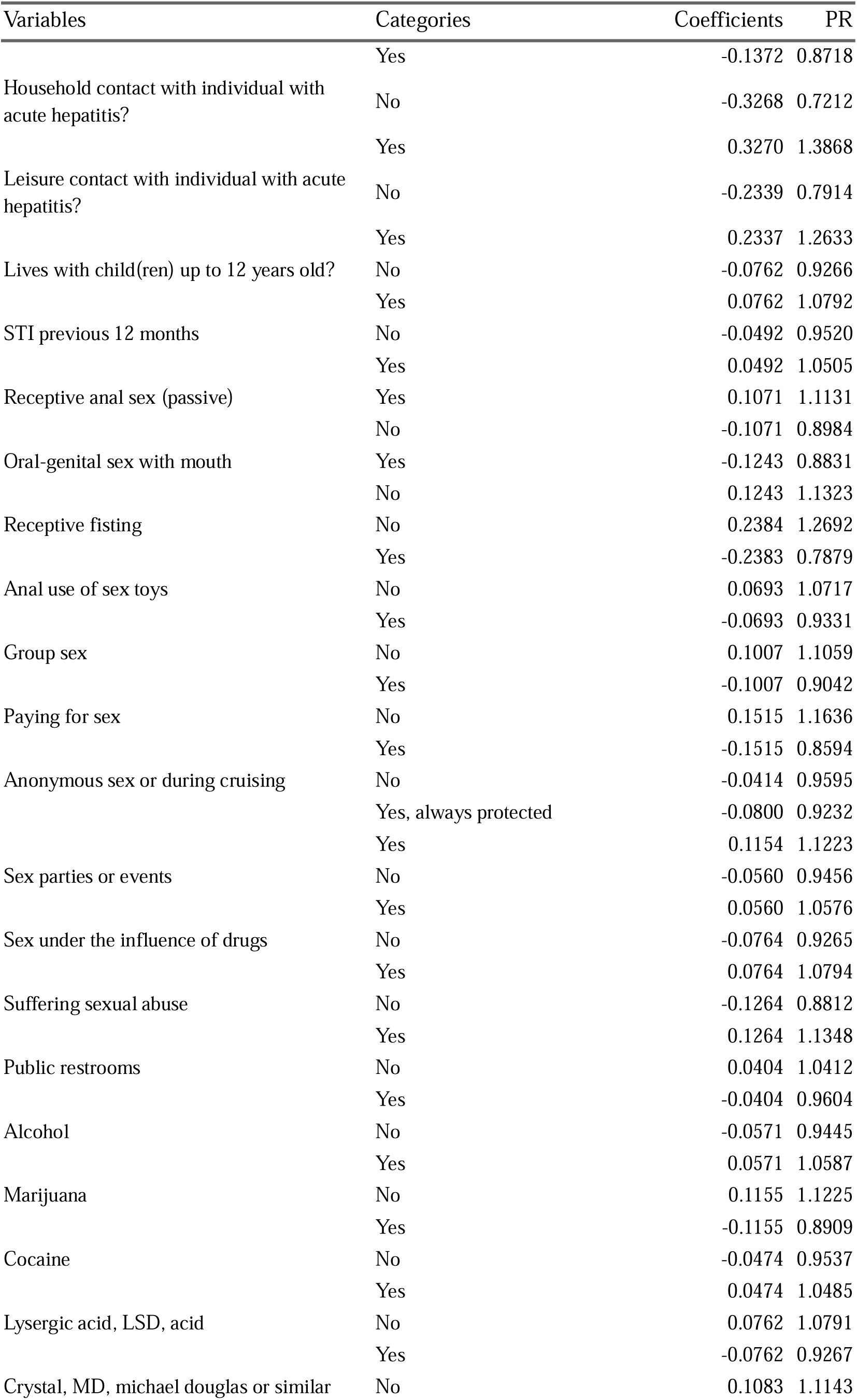
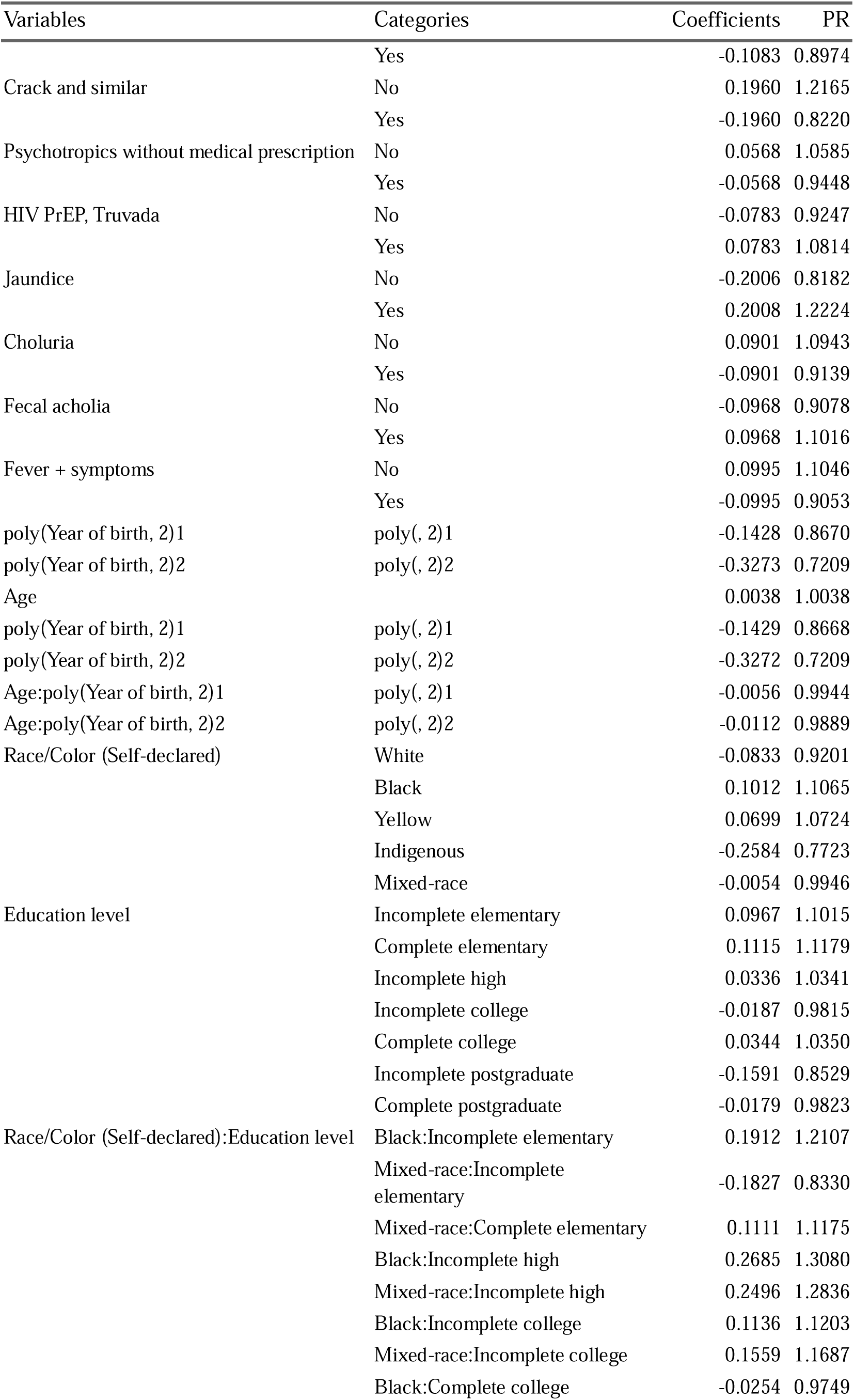
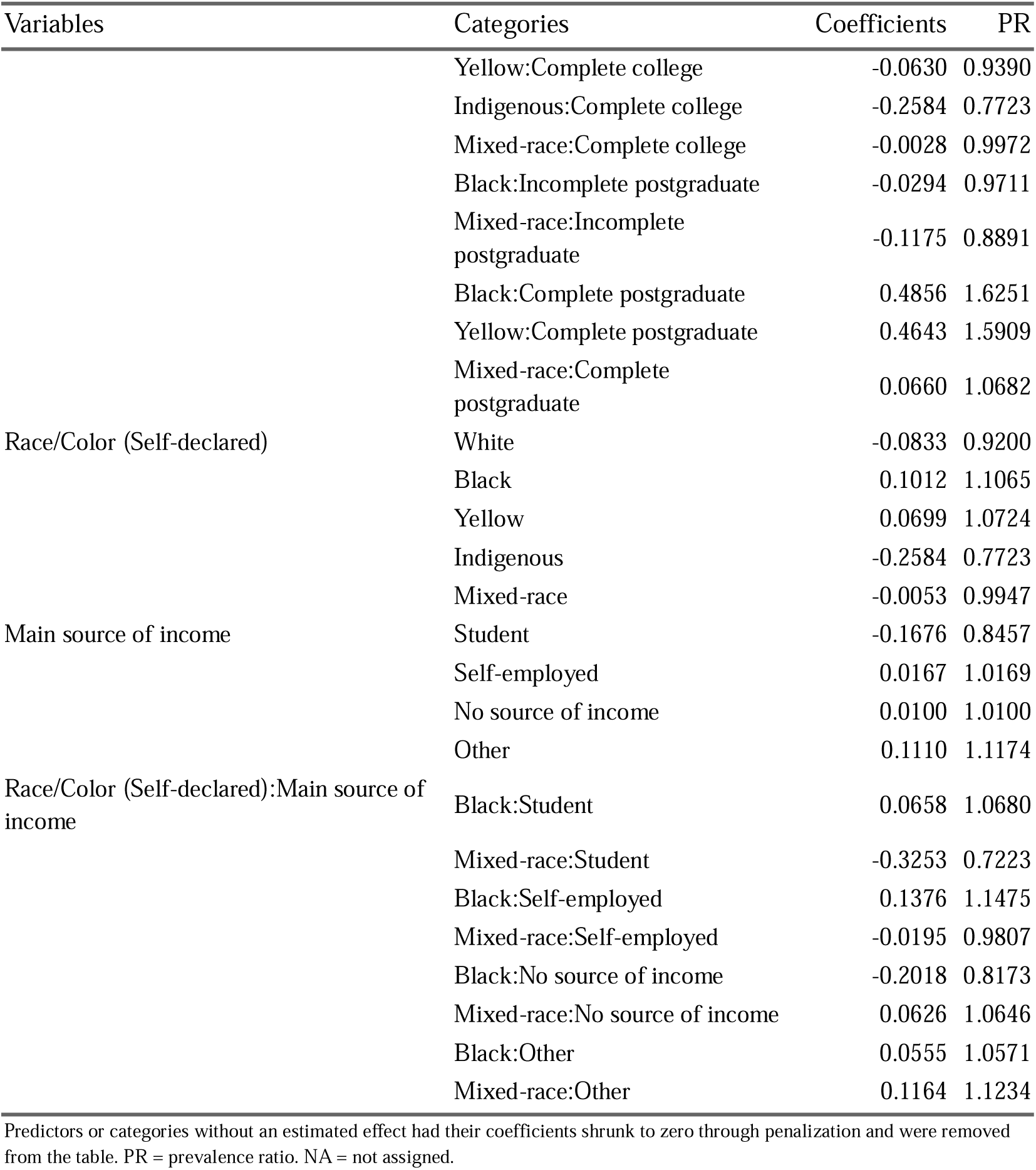
Final penalized and cross-validated Poisson model effects including place of birth and living area, and Prevalence Ratios taking HAV positive serology as outcome.

Both GLM models’ discriminations were considered good. (Figure 3 and Figure S15) The GLM model with place of birth and living place as predictors had slightly higher discrimination metrics. However, both approaches had very bad calibration metrics, showing an overestimation in the lower range of predictions and an underestimation in the higher range of predictions. Both final GLM models had better discrimination when compared to both random forest models, and they also had worse calibration when compared to the random forest models. To allow readers to estimate the risk of seropositivity among MSM, we provide a web calculator at https://pedrobrasil.shinyapps.io/INDWELL/.

**Figure 3:**
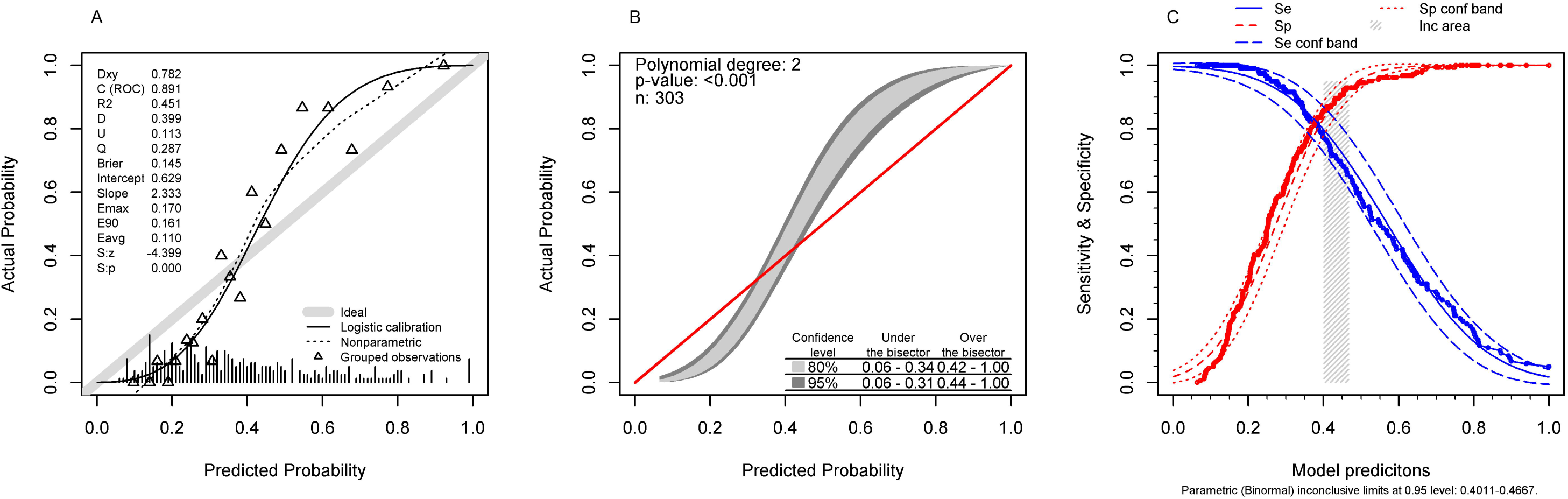

## DISCUSSION

The main results to be discussed are: (a) these findings corroborate broader trends in the shifting epidemiology of HAV in Brazil, where declining pediatric incidence has paralleled increases in adult susceptibility, especially among MSM; (b) in addition to usual social demographic exposures, there is evidence that sexual behavior and preferences, and the use of drugs also increase the risk for seropositivity among adult MSM; (c) it is possible to make reasonably accurate predictions of HAV seropositivity for the MSM population.

HAV is an intestinal infection, and it has a fecal–oral transmission pathway; therefore, its prevalence is tightly related to water and sewage access and quality. Children are usually the most exposed group to HAV infection in the presence of poor conditions. WHO characterizes patterns of HAV infection into areas of high, intermediate, or low endemicity.[24] Hepatitis A endemicity classification is determined by the seroprevalence among children aged 10-14 years: > 90% - high, 75-89% - high/medium, 60-74% - medium, 40-59% - low/medium, < 20% - low. Additionally, there is an increased risk of outbreaks as the number of susceptible adults increases, therefore, WHO also has a susceptibility classification according to adults aged 35-44 years at risk for HAV infection as follows: high: > 40%, medium: 20-39%, low/medium: 10-19%, low: 1-9%, and very low: 0%.[25] Therefore, the MSM population in our setting could be considered at high risk for HAV outbreaks.

Although the introduction of childhood vaccination into Brazil’s national program in 2014 has drastically reduced HAV incidence in children, immunity gaps persist among adults who were never exposed or vaccinated, leading to periodic outbreaks in urban centers.[18,26,27] The MSM population has the same determinants of HAV infection as the overall population, and there is evidence that the generation and cohort effects observed in general are also present in this population. This means that the generation decline in HAV seropositivity among MSM is primarily driven by a cohort effect, with younger generations having less natural immunity due to reduced childhood exposure, thereby increasing the risk of outbreaks in this population.[17,28]

Notably, our results align with those of Carvalho et al., who found a 44% seroprevalence of HAV antibodies in adults aged 18 to 40 attending the same referral center, with particularly low immunity among travelers, healthcare professionals, and household contacts of immunocompromised individuals.[29] Their predictive model highlighted age and year of birth (cohort and generation effects) as major drivers of seropositivity, with older individuals showing higher prevalence. However, unlike the broader special populations in Carvalho et al., our sample was composed exclusively of young cisgender MSM, of whom nearly 60% lacked serological evidence of prior HAV exposure despite urban residency.

In addition to sanitation-related exposures present in the overall population, in young MSM, specific sexual behaviors are associated with increased risk of HAV acquisition. The medical literature consistently identifies oro-anal contact (rimming) as an independent risk factor for HAV acquisition, regardless of sexual orientation, due to the fecal–oral route of transmission.[30] Unprotected oro-anal sex and insertive anal intercourse have both been associated with HAV infection in MSM populations.[31,32] In particular, unprotected oro-anal sex is frequently reported among MSM and is strongly linked to HAV outbreaks.[31] Insertive anal intercourse has also been independently associated with recent HAV infection in young MSM.[32]

The results in this study also show that many sexual preferences and behaviors are linked to HAV seroprevalence. However, many adjusted prevalence ratios are counterintuitive, such as PrEP and Oro-genital sex with mouth. One must consider the nature of confounding and interaction, and that some expected reasoning may not be true. For example, the PrEP itself may not increase the risk of HAV, but it may be linked to a perception of medical attention and care, and therefore, the feeling of being safe may lead to riskier behavior. It is obvious that the idea of oral-anal sex increases the exposure; however, if almost all subjects perform this behavior, it becomes a characteristic of the baseline risk of the population, and it is not possible to detect its effect within the population.

In this study, like the sex related predictors, many of the drug-related predictors had counterintuitive results. Again, one must consider that these predictors bring additional information after the usual social-demographic and sanitary exposures. It is possible that those who engage in drug use for “chemsex” are more likely to have a higher socio-economic status and are more likely to be able to protect themselves from both childhood and specific MSM exposures. Nevertheless, we found a specific increase in the adjusted prevalence ratio for those who engage in sex under the influence of drugs.

The literature adds contextual and behavioral factors that increase HAV risk in this population, including having multiple sexual partners, participation in “chemsex” (sexual activity under the influence of drugs), previous diagnosis of other sexually transmitted infections, and attendance at sex venues.[31,33,34] The use of electronic dating apps and international travel has also been implicated in facilitating the spread of HAV among MSM during recent outbreaks.[33–35] Notably, HAV outbreaks in MSM are often characterized by high rates of coinfection with other STIs and a high proportion of unvaccinated individuals.[33,34]

Molecular epidemiology studies from São Paulo have demonstrated that the majority of HAV strains identified during recent outbreaks were closely related to those circulating in European MSM outbreaks, particularly those from 2016–2017, indicating direct epidemiological links between continents.[36] Similar findings have been reported in Rio de Janeiro, where phylogenetic analyses revealed that several outbreak strains among MSM were imported from Europe and Asia, with sexual practices being the predominant risk factor for transmission.[37] Additional molecular evidence from Brazil confirms the introduction of European outbreak strains into local MSM populations, further supporting the role of international travel and interconnected sexual networks in the dissemination of HAV.[38]

In MSM, “chemsex” commonly involves the use of specific drugs, most notably methamphetamine (crystal meth), gamma-hydroxybutyrate/gamma-butyrolactone (GHB/GBL), mephedrone, cocaine, ketamine, and, to a lesser extent, 3,4-methylenedioxymethamphetamine (MDMA/ecstasy) and inhaled nitrites (“poppers”).[39–44] The core “chemsex” drugs consistently identified across European, North American, and Asian cohorts are methamphetamine, GHB/GBL, and mephedrone, with regional variations in the prevalence of ketamine, cocaine, and MDMA use.[39–44]

The association between these drugs and hepatitis A transmission was, up to this study, indirect and primarily mediated by the high-risk sexual behaviors facilitated by “chemsex”, such as condomless anal and oro-anal sex with multiple partners, group sex, and prolonged sexual sessions.[39,41,44] These behaviors increase the likelihood of fecal–oral exposure. While the medical literature robustly links “chemsex” to increased rates of sexually transmitted infections (STIs) such as HIV, syphilis, and gonorrhea, direct evidence specifically quantifying the risk of HAV transmission by individual “chemsex” drugs is limited.[39–41,43,44] However, methamphetamine, GHB/GBL, and mephedrone are most strongly associated with the “chemsex” context in which HAV outbreaks have been reported among MSM.[40–44]

There is some research using a multivariable approach that links certain predictors to HAV seropositivity among MSM, mainly age, in a variety of settings.[17,45–47] This is expected as this follows an overall trend of cumulative exposure to HAV over time. Additionally, multivariable models predicting HAV seropositivity, susceptibility, or transmission among MSM have consistently identified several predictors related to sexual behavior and sexual preferences, although the specific construct of “chemsex” is not directly included in these models. Examples of these predictors are: number of sexual partners,[32,48] type of sexual activity,[32] sexual network characteristics,[47] sexual preferences,[47] and other risk behaviors such as sharing needles without cleaning them.[32]

Additionally, the literature notes that MSM with high-risk sexual behaviors—such as sex with multiple partners, use of electronic dating apps, and attendance at sex venues— are disproportionately affected during HAV outbreaks, although these factors are more often described in outbreak reports and epidemiologic summaries than as formal predictors in multivariable models.[34] However, for predictive individual risk purposes, the performance of these models is limited, and it was either not informed or considered poor.

This result adds to the literature an individual prediction tool with reasonable accuracy, specifically for the MSM population. This model includes social-demographic characteristics (including the cohort effect and interaction of race, education and source of income), sexual preferences and behaviors, and several drugs representing the “chemsex” construct. The added value of our study lies in the primary data collection among a highly specific and underrepresented risk group.

In summary, we advocate for urgent policy adjustments to expand adult HAV vaccination strategies in Brazil. This is especially warranted given the high susceptibility of HAV within this population, the predictable recurrence of outbreaks, and the socio-structural vulnerabilities of MSM communities. Future efforts should also investigate barriers to adult vaccination uptake, as well as the acceptability and feasibility of targeted immunization in sexual health services.

## CONCLUSION

This study reveals a high susceptibility of cisgender MSM aged 18 to 35 in Rio de Janeiro to HAV infection, particularly among black individuals, with lower education, unemployed / students, with unprotected and promiscuous sex practices, and engaging in “chemsex”. These findings highlight the enduring impact of socioeconomic inequities on enteric virus exposure and underscore the need for targeted public health strategies. Given the frequency of severe HAV cases among adults and the HAV susceptibility in MSM, catch-up vaccination efforts, health education initiatives tailored to vulnerable populations, and improved surveillance of HAV circulation in urban MSM communities are warranted. One may use the prediction model to estimate the risk of seropositivity/susceptibility to HAV in aid of deciding either to perform a HAV test or to vaccinate against HAV, or even to estimate HAV seroprevalence in MSM populations where the HAV test is not easily available. Integrating these approaches into broader sexual health services could enhance immunity coverage and reduce the burden of preventable infections in key populations.

## Data Availability

All data produced in the present study are available upon reasonable request to the authors

## AUTHORS CONTRIBUTIONS

Alberto dos Santos de Lemos – Conceptualization, Funding acquisition, Data curation, Investigation, Roles/Writing - original draft

Luciana Gomes Pedro Brandão – Investigation, Writing - review & editing

Marcellus Dias da Costa – Conceptualization, Investigation, Writing - review & editing

Sergio Carlos Assis de Jesus Jr -Project administration

Daniel Marinho da Costa -Project administration

Ananza Taina da Silva Villaça -Investigation

Flavio de Carvalho -Investigation

Margareth Catoia Varela - Data curation, Writing - review & editing

Pedro Emmanuel Alvarenga Americano do Brasil -Methodology, Visualization, Roles/Writing - original draft, Writing - review & editing

## AKNOWLEDGEMENTS

A research grant from the Investigator-Initiated Studies Program of Merck Sharp & Dohme LLC partially supported this research. The opinions expressed in this paper are those of the authors and do not necessarily represent those of Merck Sharp & Dohme LLC.

## DECLARATION OF GENERATIVE AI AND AI-ASSISTED TECHNOLOGIES IN THE WRITING PROCESS

During the preparation of this work, the author(s) used Grammarly to check grammar, spelling, and increase reading fluidity. After using this tool/service, the author(s) reviewed and edited the content as needed and take full responsibility for the content of the publication.

## AUTHORSHIP

All authors attest they meet the ICMJE criteria for authorship.

