## supplemental file for "Seroprevalence and predictors of Hepatitis A virus immunity among young MSM in urban Brazil: a cross-sectional study at a referral center"

Additional results HAV among MSM

22/08/2025 12:10

### 1. Missing data assessment

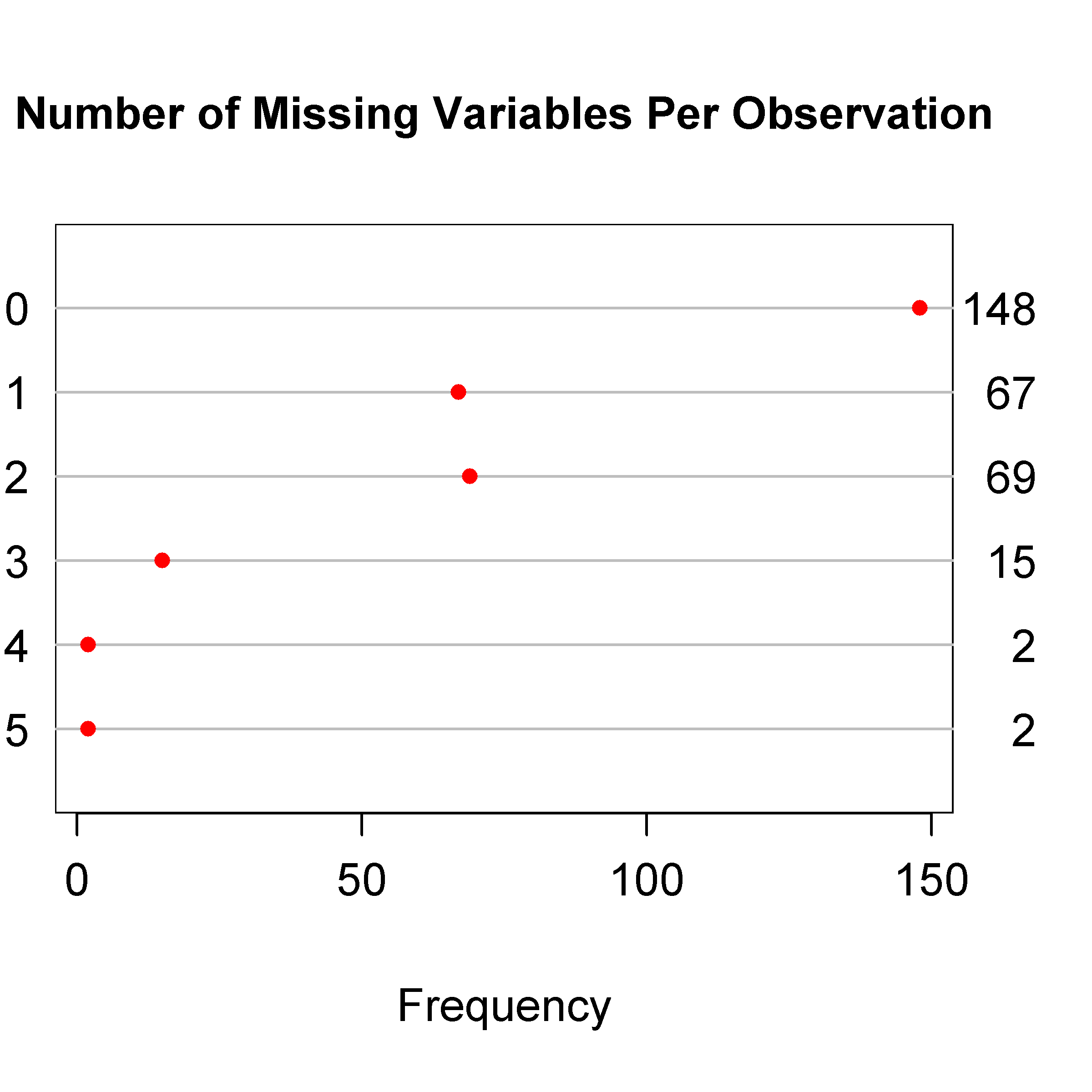

**Figure S****1:** Missing data assessment per observation for the socio-demographic and potential exposure characteristics

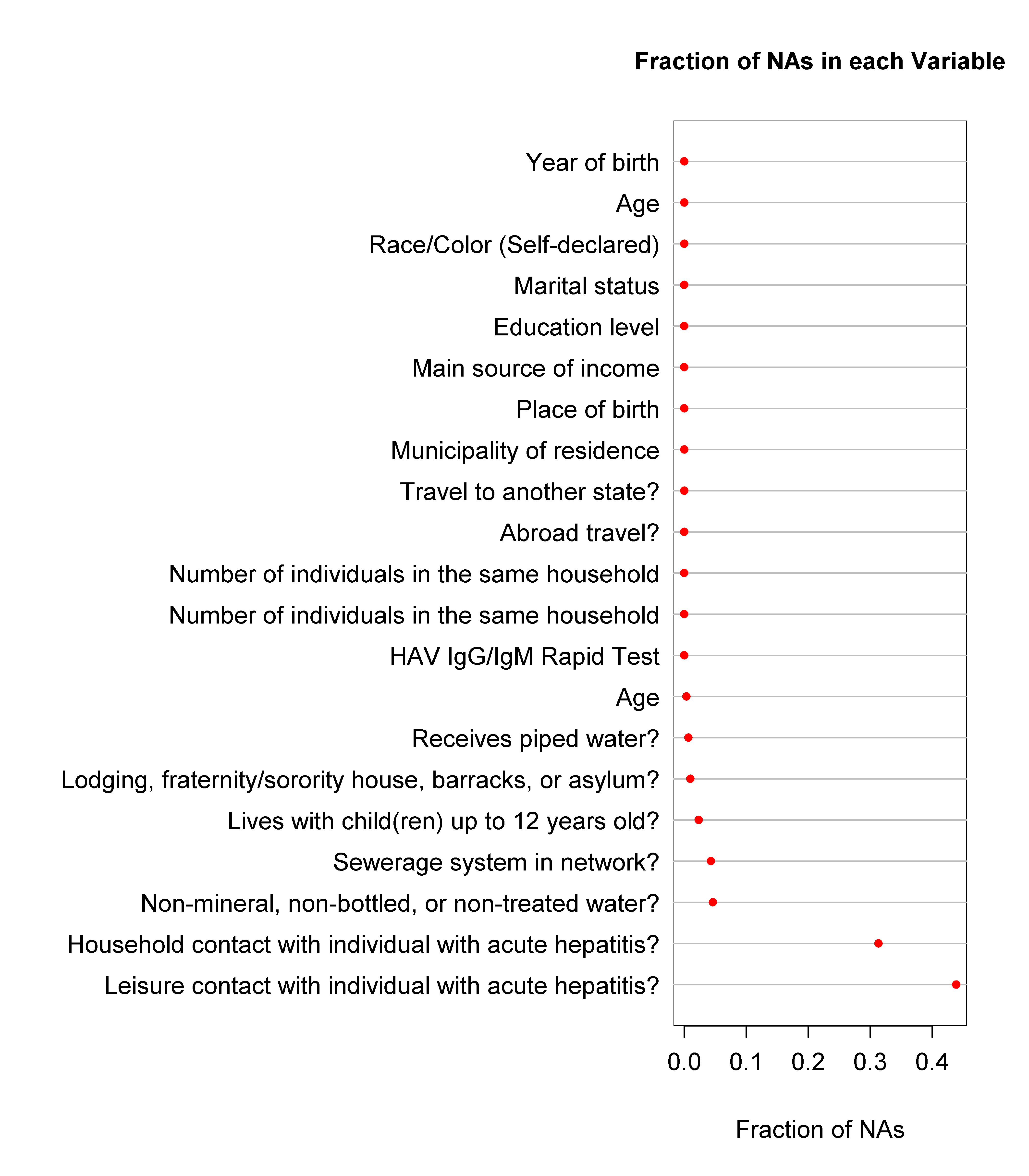

**Figure S****2:** Missing data assessment per variable for the socio-demographic and potential exposure characteristics

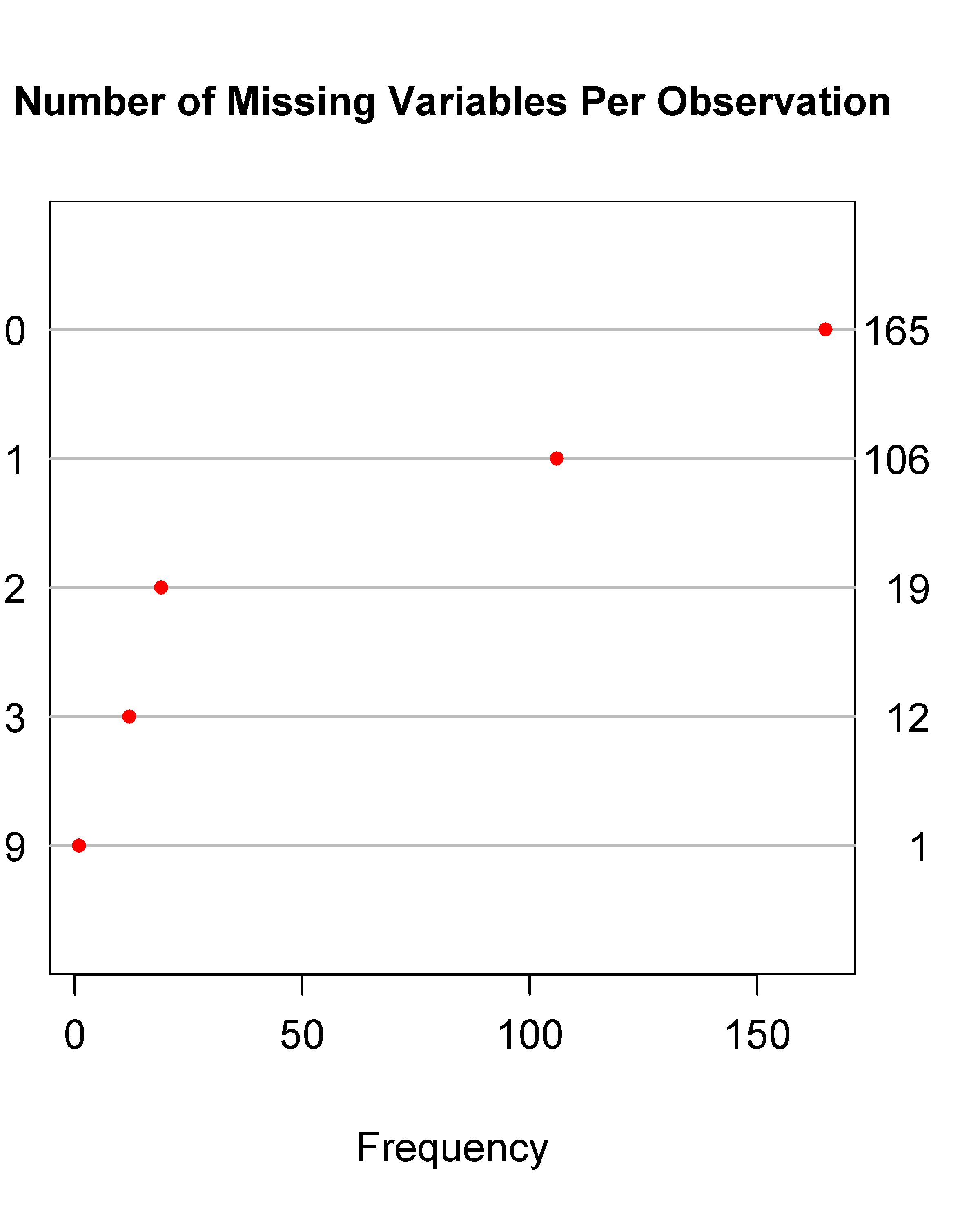

**Figure S****3:** Missing data assessment per observation for the sexual preferences and behaviors.

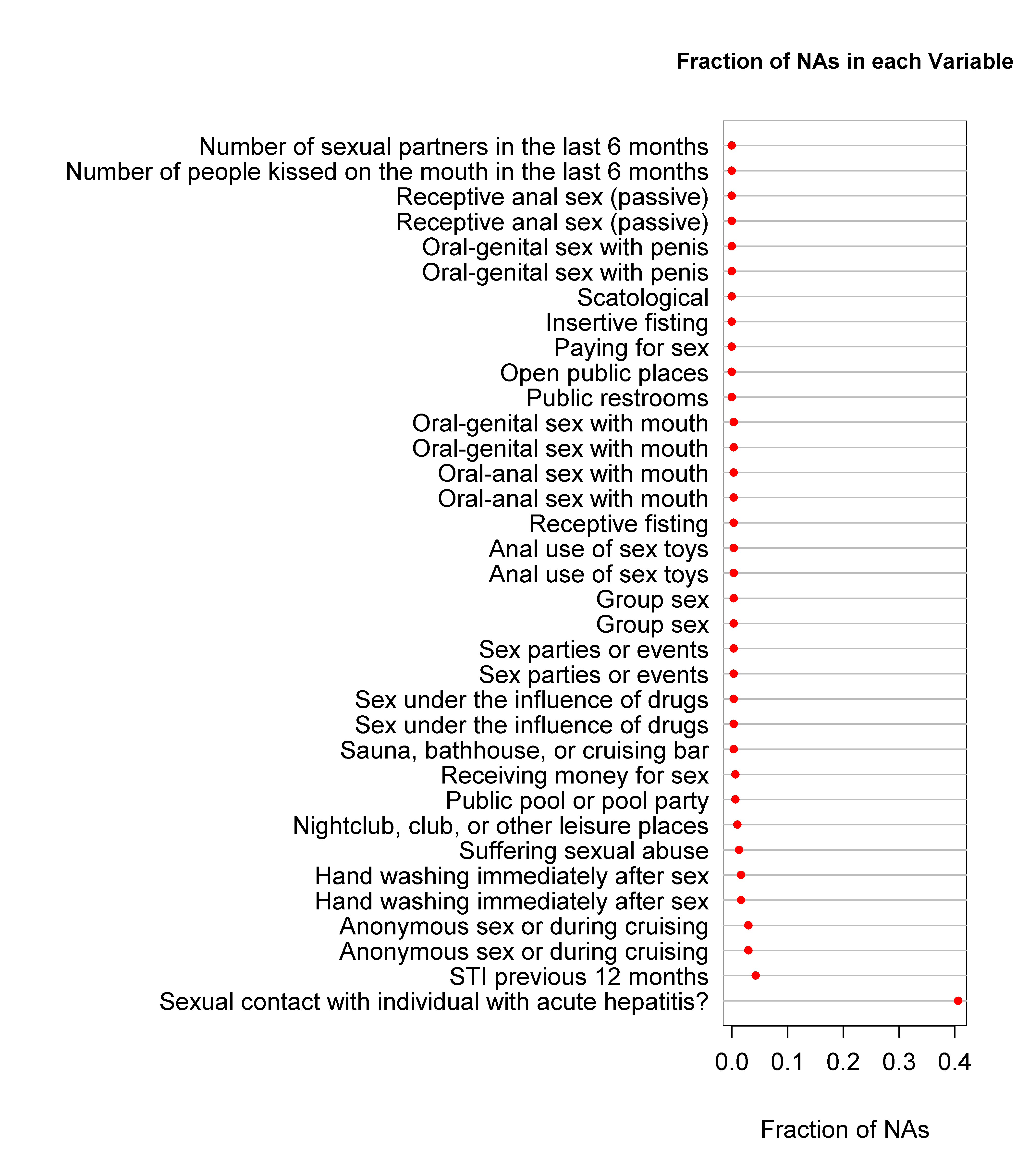

**Figure S****4:** Missing data assessment per variable for the sexual preferences and behaviors

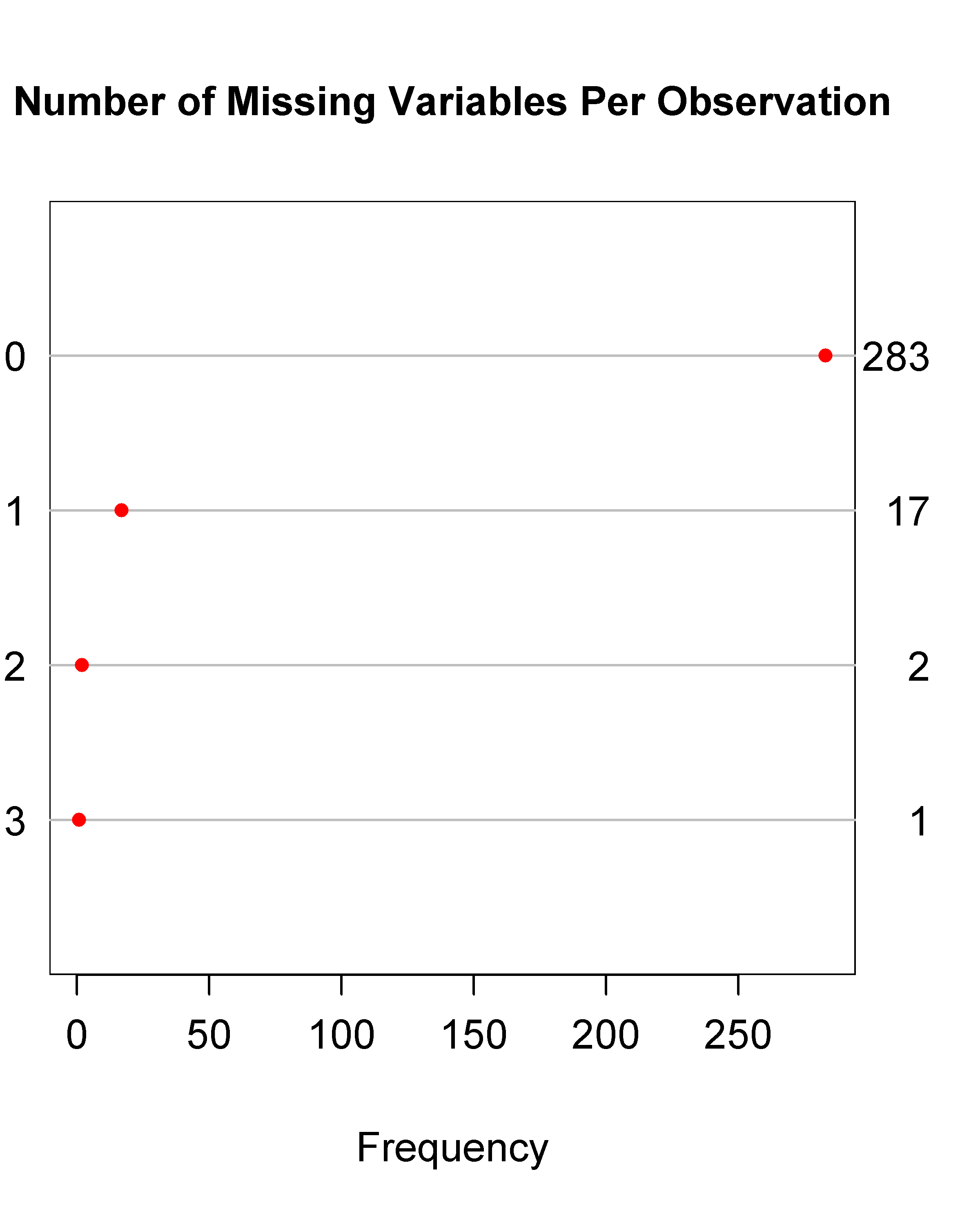

**Figure S****5:** Missing data assessment per observation for the drug use.

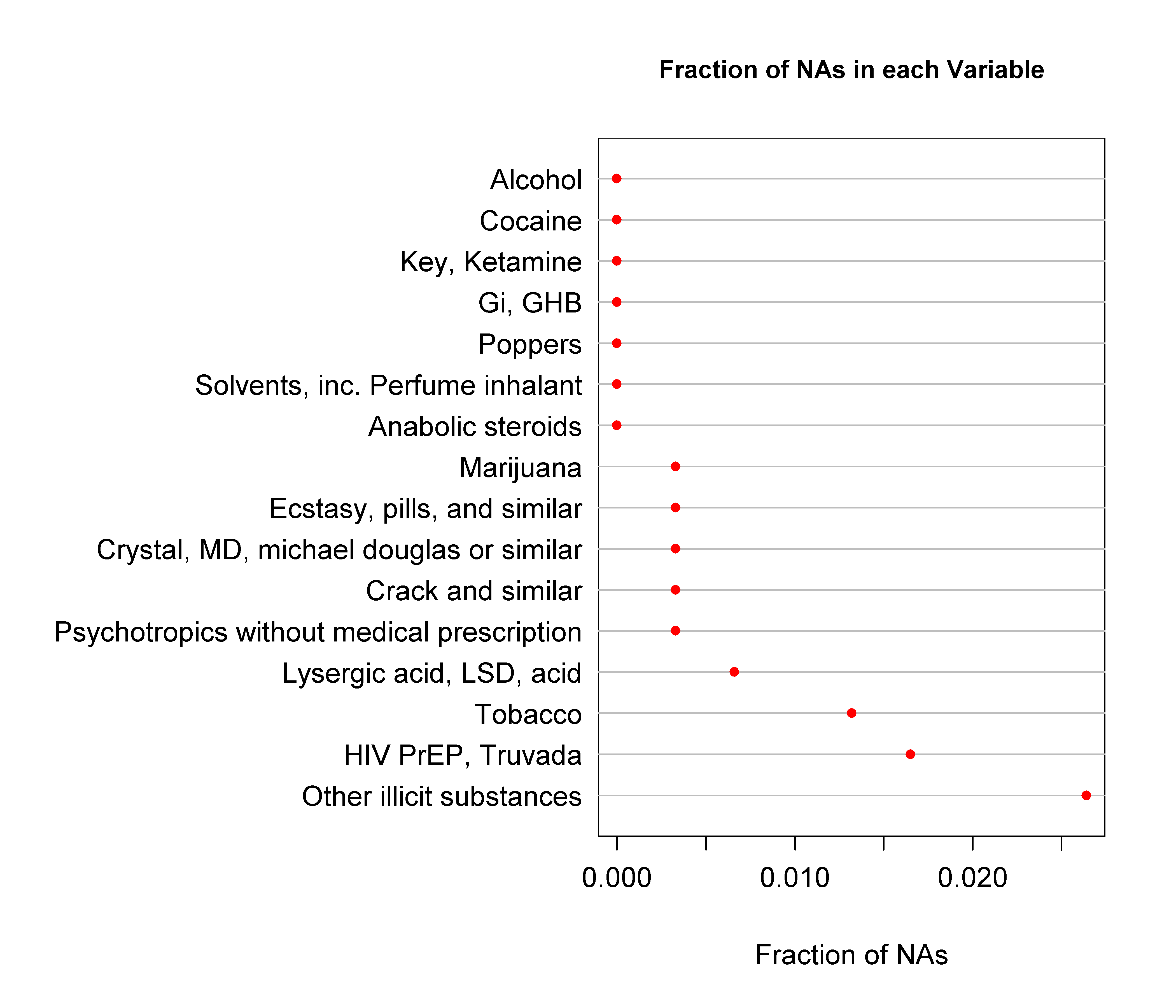

**Figure S****6:** Missing data assessment per variable for the drug use.

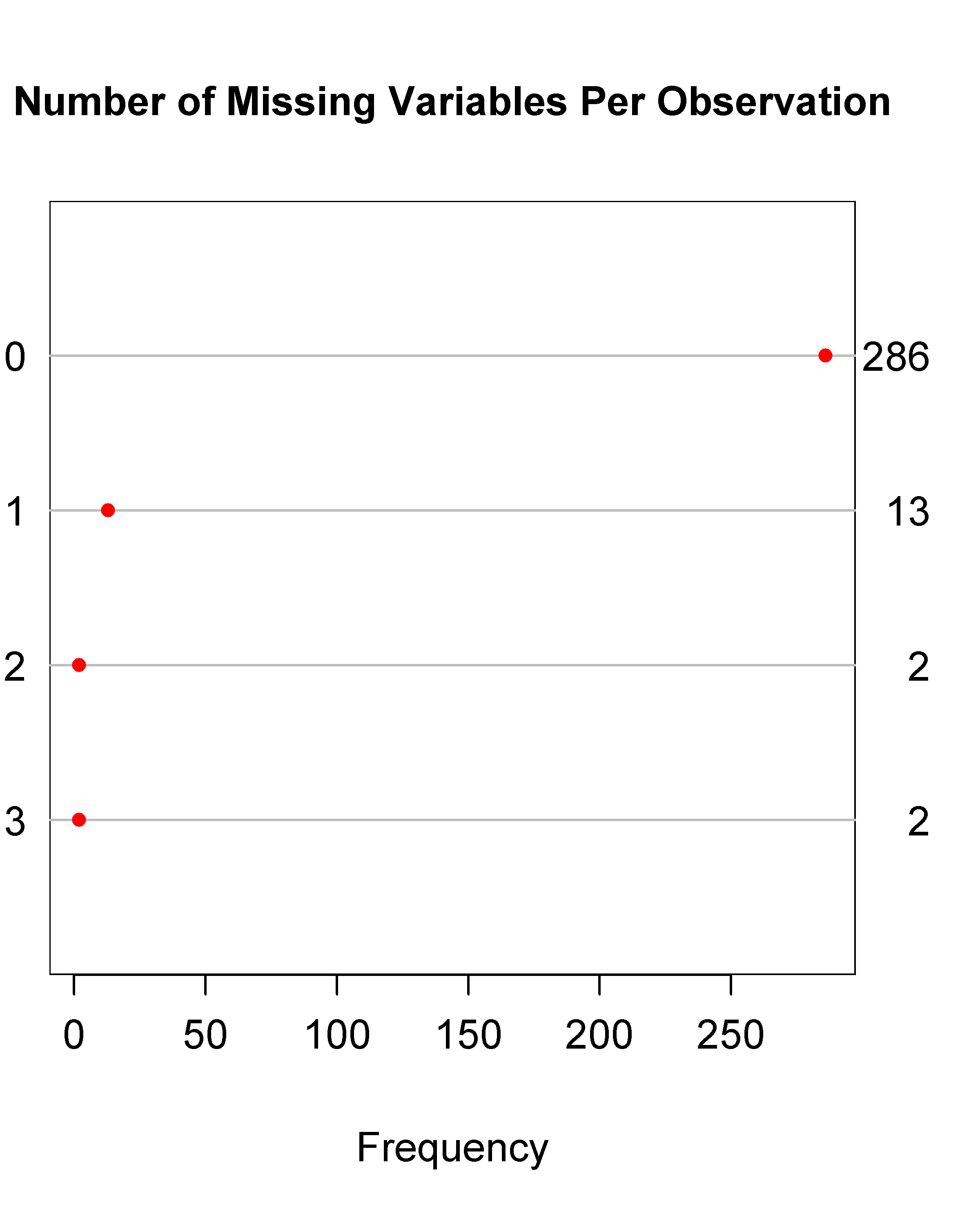

**Figure S****7:** Missing data assessment per observation for signs and symptoms.

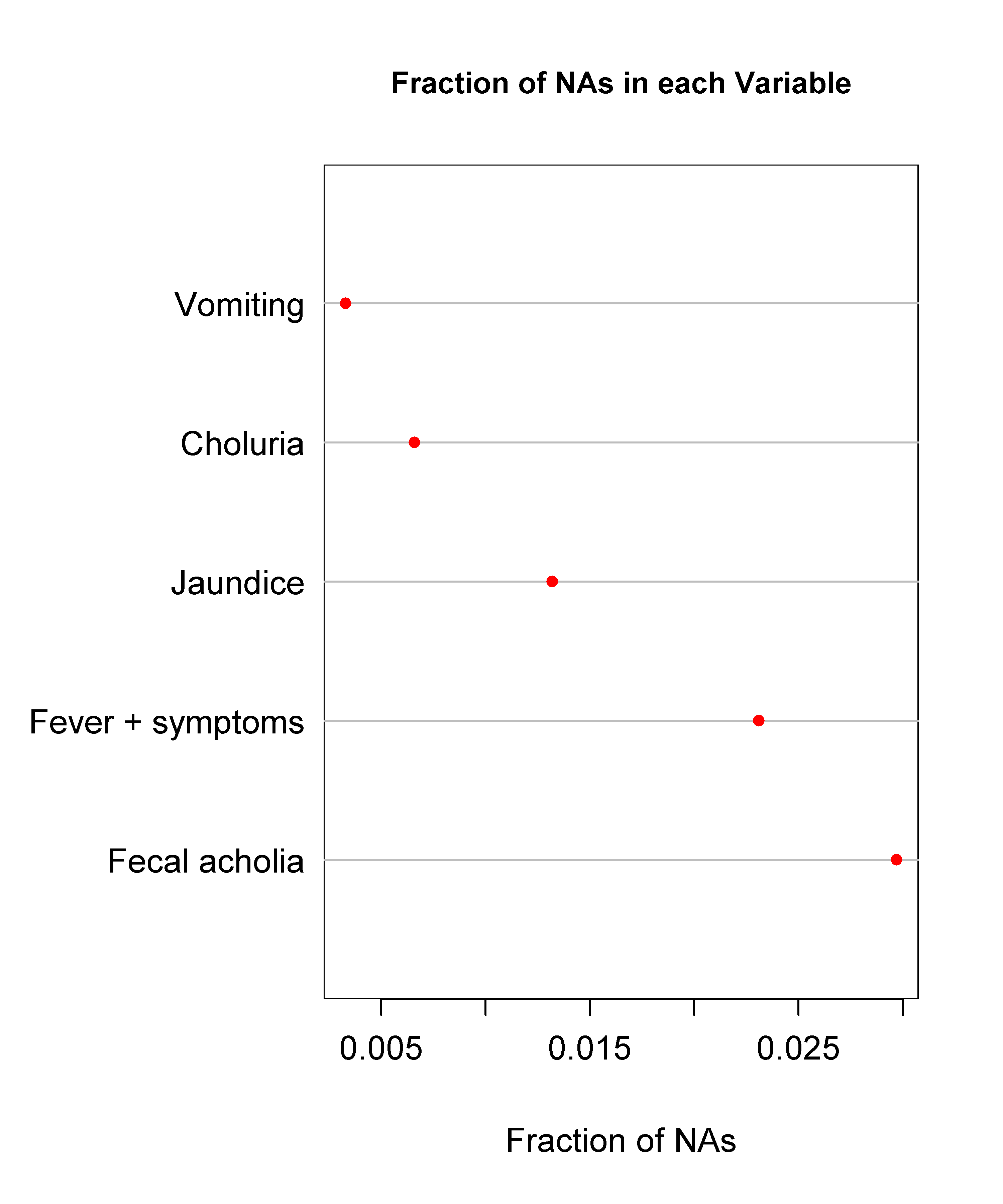

**Figure S****8:** Missing data assessment per variable for signs and symptoms.

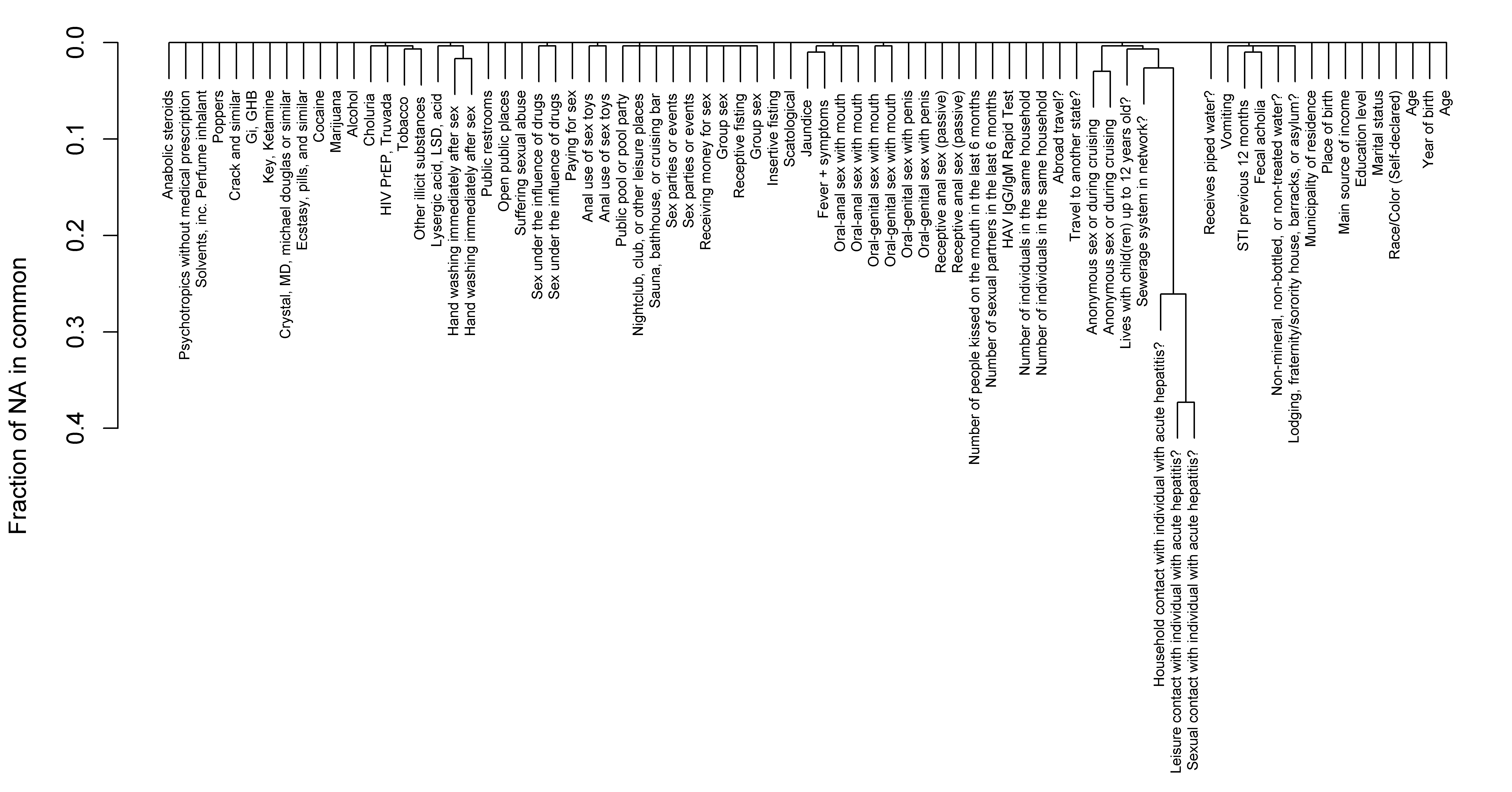

**Figure S****9:** Clustered missing data assessment for all variables.

### 2. Random forest additional results.

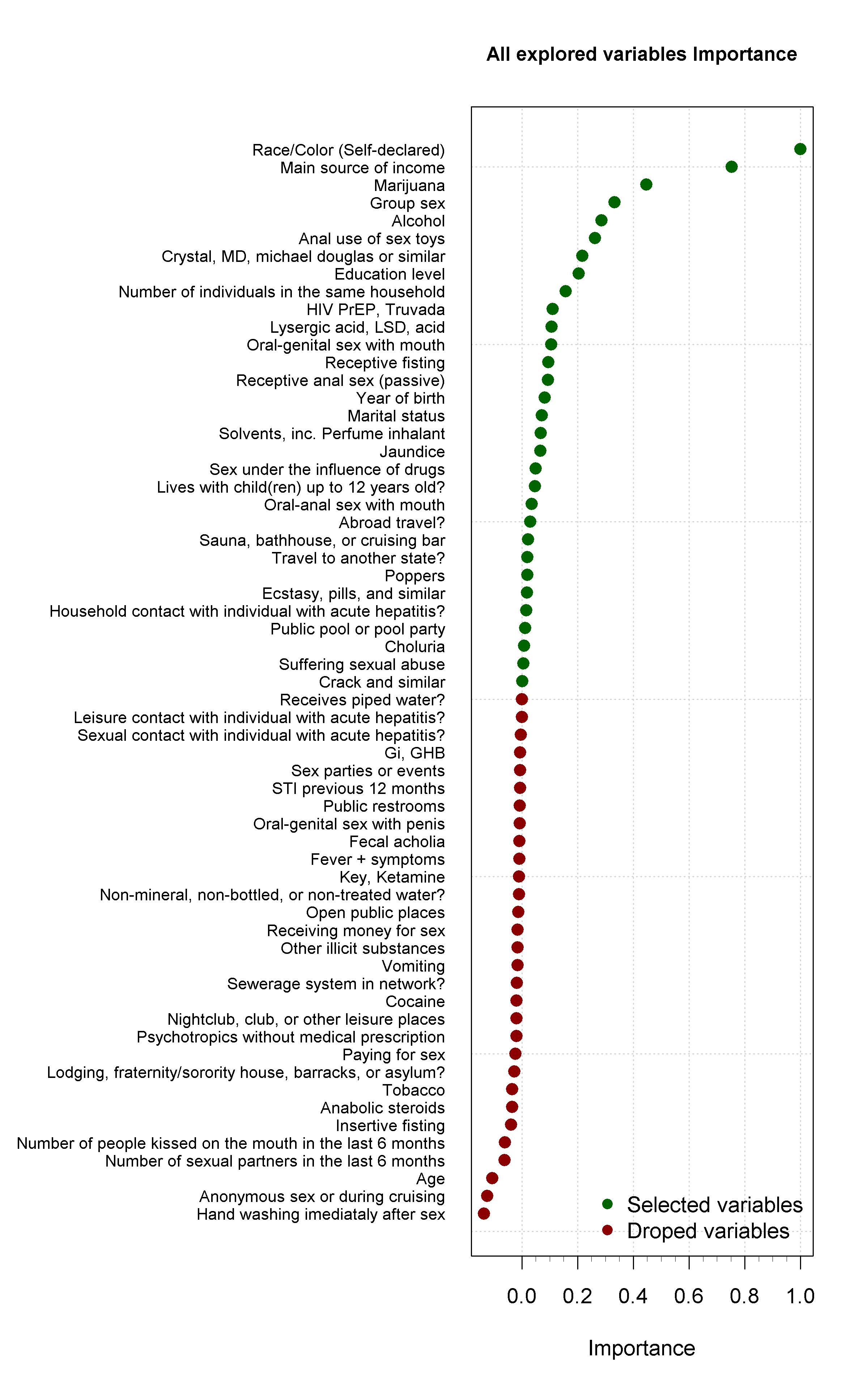

**Figure S****10:** Overall importance of all explored predictors in the random forest approach without the place of birth and living place variables.

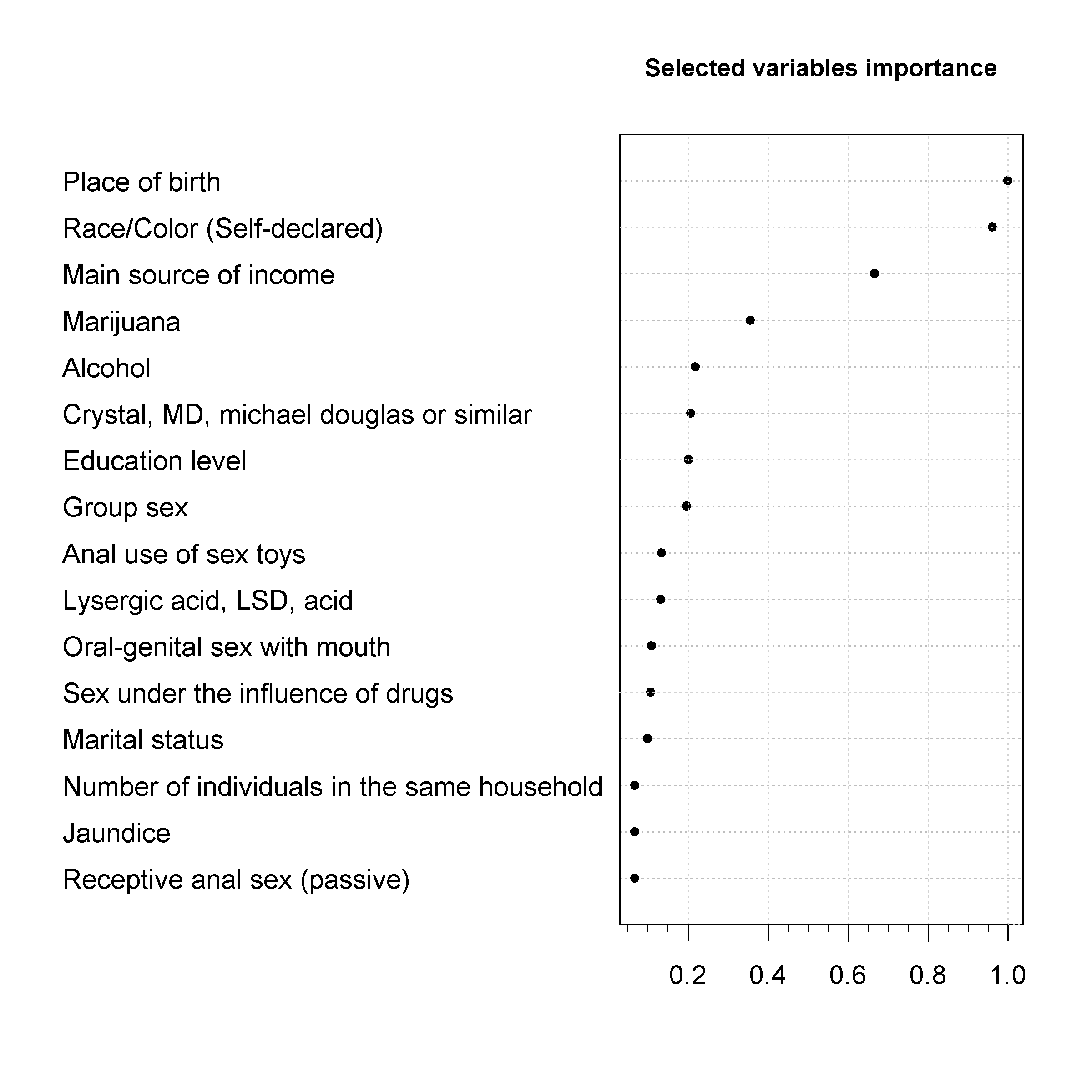

**Figure S****11:** Importance of the selected predictors in the random forest approach with the place of birth and living place variables.

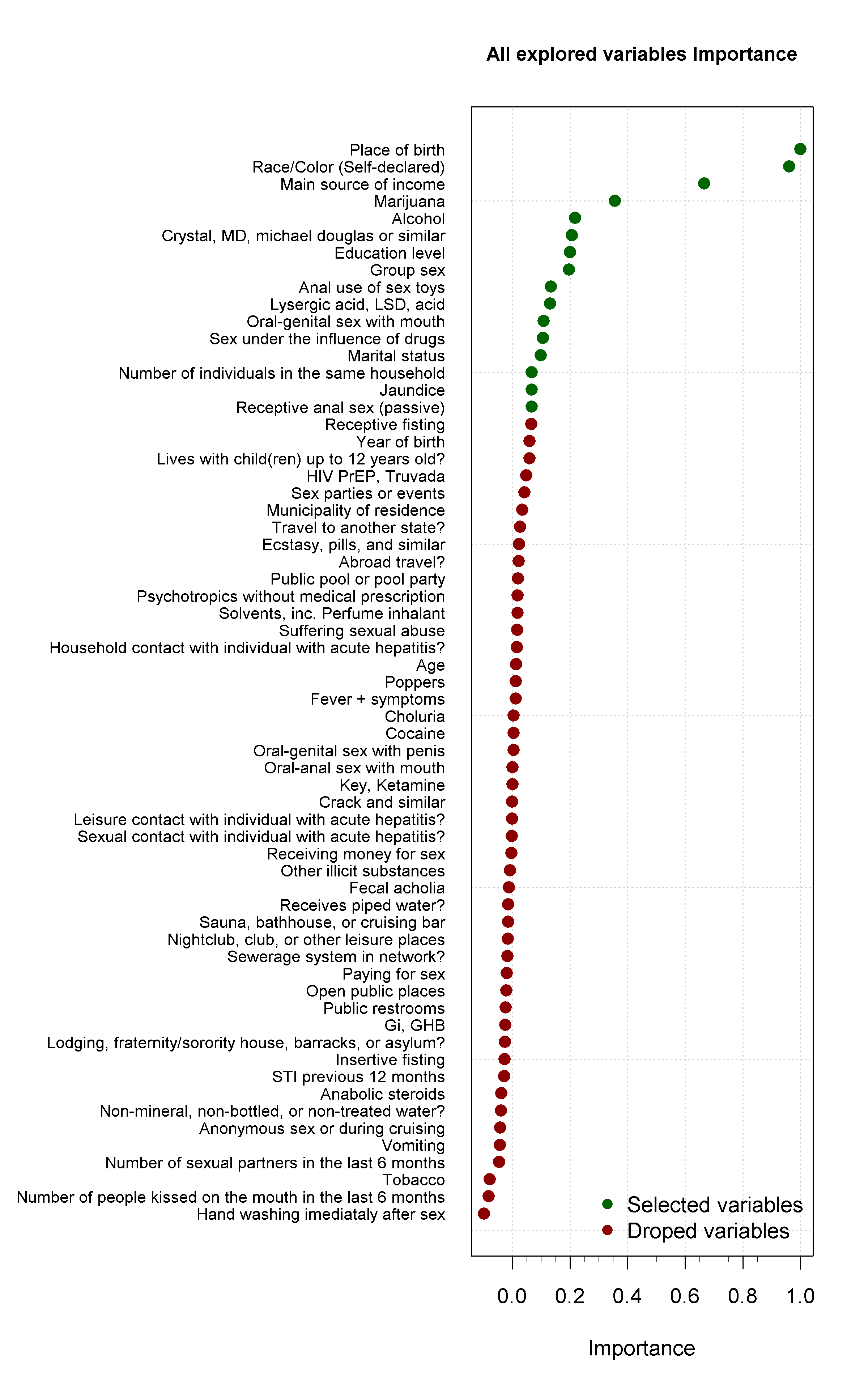

**Figure S****12:** Overall importance of all explored predictors in the random forest approach with the place of birth and living place variables.

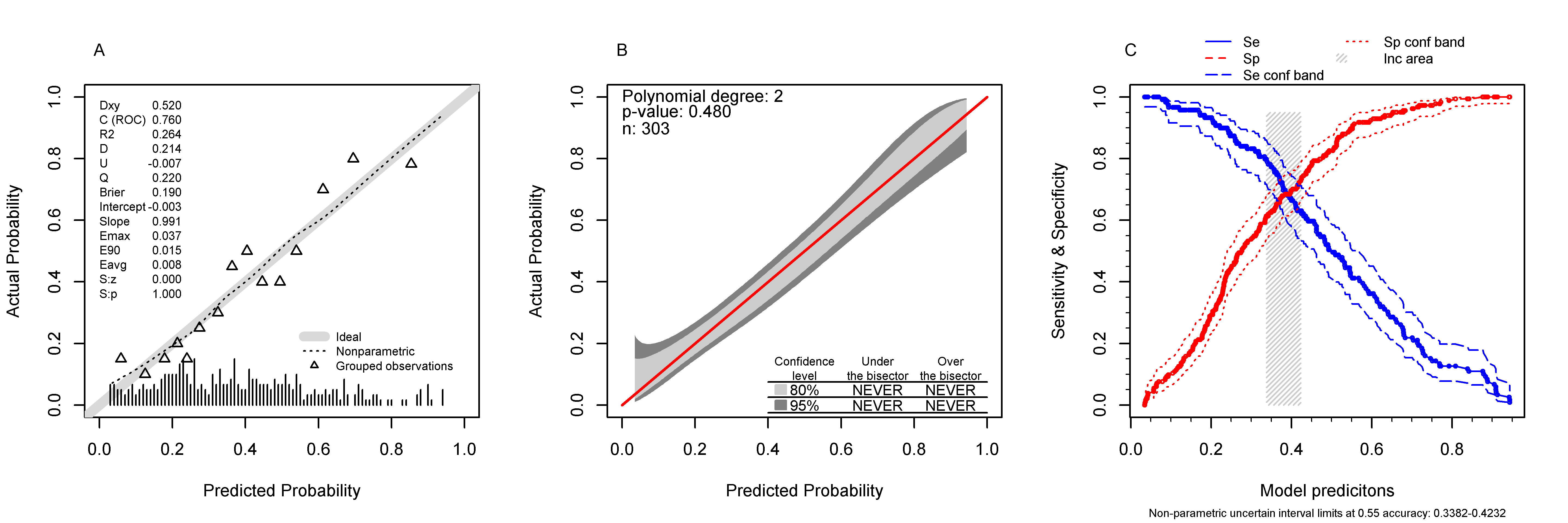

**Figure S****13:** Random forest discrimination and calibration performance including the place of birth as a predictor variable.

**Table S1:** Random Forest Regression Model Cross-validation metrics.

| Metric | Value |
| --- | --- |
| Fit MSE | 0.1902 |
| Fit % Variance Explained | 20.26 |
| Median Permuted MSE | 0.1018 |
| Median Permuted % Variance Explained | 57.23 |
| Median Cross-validation RMSE | 0.4458 |
| Median Cross-validation MBE | -0.0052 |
| Median Cross-validation MAE | 0.3837 |
| Range of Kolmogorov-Smirnov p-values | 3.467e-54 - 1.998e-15 |
| Range of Kolmogorov-Smirnov D statistic | 0.5048 - 0.7368 |
| RMSE Cross-validation Error Variance | 0.000313 |
| MBE Cross-validation Error Variance | 0.002953 |
| MAE Cross-validation Error Variance | 0.000224 |
| MSE = mean squared error; RMSE = Root Mean Squared Error; MBE = Mean Bias Error; MAE = Mean Absolute Error. | |

### 3. GLM additional results

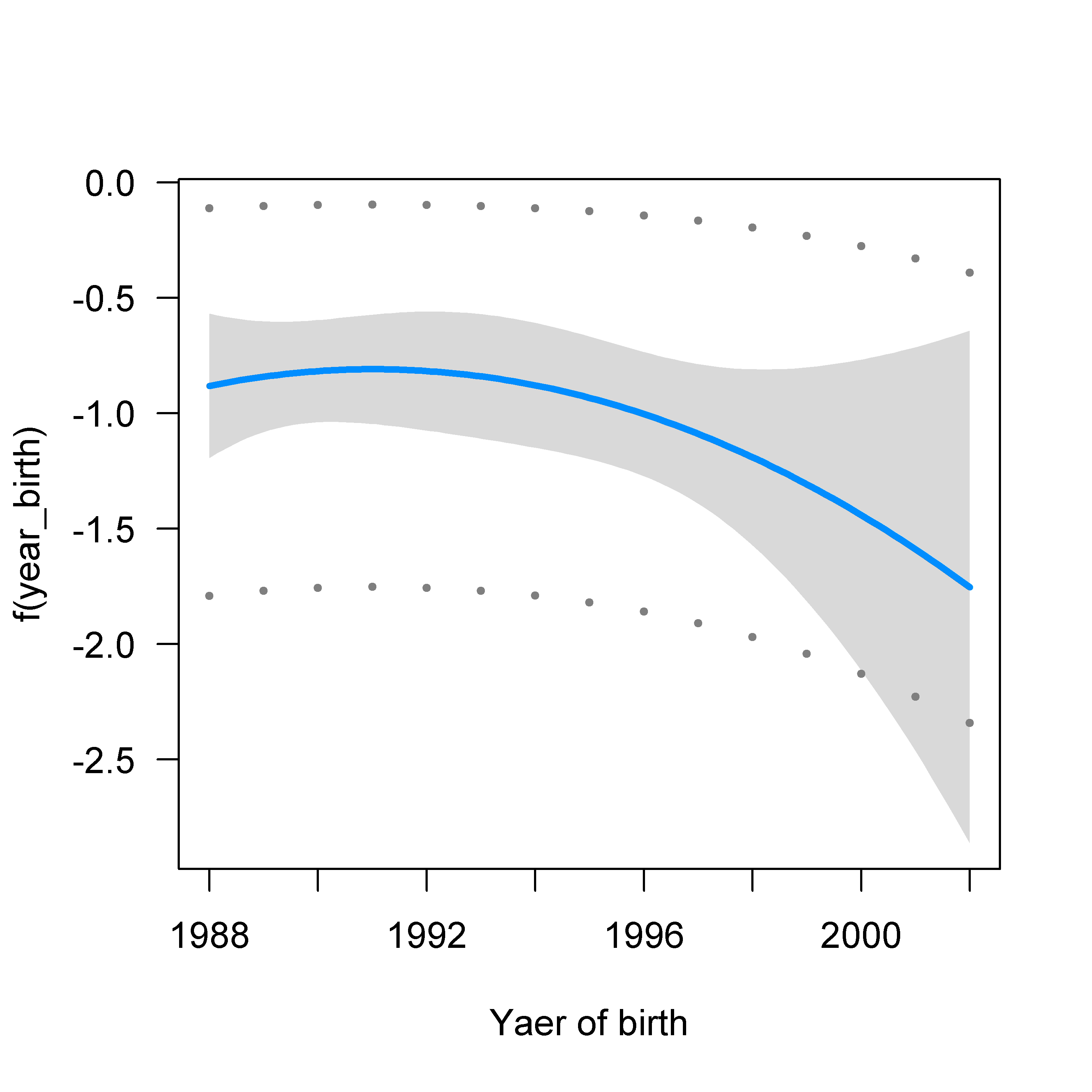

**Figure S****14:** Visualization of the Year of birth non-linear relationship (polynomial with 2 degrees) with the Hepatitis A outcome in a Poisson general linear model

**Table S****2:** Final penalized and cross-validated Poisson general linear model effects and Prevalence Ratios taking HAV positive serology as outcome.

| Variables | Categories | Coefficients | PR |
| --- | --- | --- | --- |
| (Intercept) | - | -1.1886 | 0.3047 |
| Age |  | 0.0038 | 1.0038 |
| Race/Color (Self-declared) | White | -0.0839 | 0.9195 |
|  | Black | 0.0975 | 1.1024 |
|  | Yellow | 0.0764 | 1.0794 |
|  | Indigenous | -0.2819 | 0.7543 |
|  | Mixed-race | -0.0013 | 0.9987 |
| Marital status | Single | 0.0503 | 1.0516 |
|  | Married | -0.0447 | 0.9563 |
|  | Widowed | 0.1267 | 1.1351 |
|  | Separated | -0.3947 | 0.6739 |
| Education level | Complete high | -0.0078 | 0.9923 |
|  | Incomplete elementary | 0.1044 | 1.1101 |
|  | Complete elementary | 0.1807 | 1.1980 |
|  | Incomplete high | 0.0599 | 1.0618 |
|  | Incomplete college | -0.0250 | 0.9753 |
|  | Complete college | 0.0354 | 1.0361 |
|  | Incomplete postgraduate | -0.1741 | 0.8402 |
|  | Complete postgraduate | -0.0011 | 0.9989 |
| Main source of income | Employed | 0.0619 | 1.0639 |
|  | Student | -0.1733 | 0.8409 |
|  | Self-employed | 0.0135 | 1.0136 |
|  | No source of income | 0.0031 | 1.0031 |
|  | Other | 0.1086 | 1.1147 |
| Non-mineral, non-bottled, or non-treated water? | No | -0.0372 | 0.9635 |
|  | Yes | 0.0372 | 1.0379 |
| Receives piped water? | No | 0.0783 | 1.0815 |
|  | Yes | -0.0784 | 0.9246 |
| Travel to another state? | No | 0.0433 | 1.0443 |
|  | Yes | -0.0434 | 0.9576 |
| Abroad travel? | No | 0.1550 | 1.1676 |
|  | Yes | -0.1550 | 0.8564 |
| Household contact with individual with acute hepatitis? | No | -0.3090 | 0.7342 |
|  | Yes | 0.3092 | 1.3624 |
| Leisure contact with individual with acute hepatitis? | No | -0.2604 | 0.7707 |
|  | Yes | 0.2602 | 1.2972 |
| Lives with child(ren) up to 12 years old? | No | -0.0705 | 0.9319 |
|  | Yes | 0.0705 | 1.0731 |
| STI previous 12 months | No | -0.0480 | 0.9531 |
|  | Yes | 0.0480 | 1.0492 |
| Receptive anal sex (passive) | Yes | 0.1123 | 1.1189 |
|  | No | -0.1123 | 0.8937 |
| Oral-genital sex with penis | Yes | -0.0331 | 0.9675 |
|  | No | 0.0330 | 1.0336 |
| Oral-genital sex with mouth | Yes | -0.1433 | 0.8665 |
|  | No | 0.1433 | 1.1541 |
| Receptive fisting | No | 0.2526 | 1.2874 |
|  | Yes | -0.2526 | 0.7768 |
| Anal use of sex toys | No | 0.0802 | 1.0835 |
|  | Yes | -0.0802 | 0.9229 |
| Group sex | No | 0.1162 | 1.1232 |
|  | Yes | -0.1162 | 0.8903 |
| Paying for sex | No | 0.1341 | 1.1435 |
|  | Yes | -0.1341 | 0.8745 |
| Anonymous sex or during cruising | No | -0.0491 | 0.9521 |
|  | Yes, always protected | -0.0809 | 0.9223 |
|  | Yes | 0.1266 | 1.1350 |
| Sex parties or events | No | -0.0533 | 0.9481 |
|  | Yes | 0.0533 | 1.0547 |
| Sex under the influence of drugs | No | -0.0860 | 0.9176 |
|  | Yes | 0.0860 | 1.0898 |
| Suffering sexual abuse | No | -0.1263 | 0.8814 |
|  | Yes | 0.1263 | 1.1346 |
| Public restrooms | No | 0.0499 | 1.0512 |
|  | Yes | -0.0499 | 0.9513 |
| Alcohol | No | -0.0481 | 0.9530 |
|  | Yes | 0.0481 | 1.0493 |
| Marijuana | No | 0.1185 | 1.1258 |
|  | Yes | -0.1185 | 0.8883 |
| Cocaine | No | -0.0715 | 0.9310 |
|  | Yes | 0.0715 | 1.0741 |
| Lysergic acid, LSD, acid | No | 0.0763 | 1.0793 |
|  | Yes | -0.0763 | 0.9265 |
| Crystal, MD, michael douglas or similar | No | 0.1223 | 1.1301 |
|  | Yes | -0.1223 | 0.8849 |
| Crack and similar | No | 0.2044 | 1.2268 |
|  | Yes | -0.2044 | 0.8151 |
| Psychotropics without medical prescription | No | 0.0696 | 1.0721 |
|  | Yes | -0.0696 | 0.9328 |
| HIV PrEP, Truvada | No | -0.0943 | 0.9100 |
|  | Yes | 0.0943 | 1.0988 |
| Jaundice | No | -0.2091 | 0.8113 |
|  | Yes | 0.2093 | 1.2328 |
| Choluria | No | 0.1151 | 1.1219 |
|  | Yes | -0.1151 | 0.8913 |
| Fecal acholia | No | -0.0892 | 0.9147 |
|  | Yes | 0.0892 | 1.0933 |
| Fever + symptoms | No | 0.1160 | 1.1230 |
|  | Yes | -0.1160 | 0.8905 |
| poly(Year of birth, 2)1 | poly(, 2)1 | -0.1841 | 0.8318 |
| poly(Year of birth, 2)2 | poly(, 2)2 | -0.3863 | 0.6795 |
| Age |  | 0.0038 | 1.0038 |
| poly(Year of birth, 2)1 | poly(, 2)1 | -0.1844 | 0.8316 |
| poly(Year of birth, 2)2 | poly(, 2)2 | -0.3863 | 0.6796 |
| Age:poly(Year of birth, 2)1 | poly(, 2)1 | -0.0066 | 0.9934 |
| Age:poly(Year of birth, 2)2 | poly(, 2)2 | -0.0137 | 0.9864 |
| Race/Color (Self-declared) | White | -0.0839 | 0.9195 |
|  | Black | 0.0976 | 1.1025 |
|  | Yellow | 0.0763 | 1.0793 |
|  | Indigenous | -0.2819 | 0.7543 |
|  | Mixed-race | -0.0012 | 0.9988 |
| Education level | Incomplete elementary | 0.1046 | 1.1102 |
|  | Complete elementary | 0.1813 | 1.1988 |
|  | Incomplete high | 0.0601 | 1.0620 |
|  | Incomplete college | -0.0250 | 0.9753 |
|  | Complete college | 0.0354 | 1.0360 |
|  | Incomplete postgraduate | -0.1741 | 0.8402 |
|  | Complete postgraduate | -0.0012 | 0.9988 |
| Race/Color (Self-declared):Education level | Black:Incomplete elementary | 0.2094 | 1.2330 |
|  | Mixed-race:Incomplete elementary | -0.1866 | 0.8298 |
|  | Mixed-race:Complete elementary | 0.1810 | 1.1984 |
|  | Black:Incomplete high | 0.2810 | 1.3244 |
|  | Mixed-race:Incomplete high | 0.2864 | 1.3316 |
|  | Black:Incomplete college | 0.1425 | 1.1532 |
|  | Mixed-race:Incomplete college | 0.1559 | 1.1687 |
|  | Black:Complete college | -0.0229 | 0.9774 |
|  | Yellow:Complete college | -0.0494 | 0.9518 |
|  | Indigenous:Complete college | -0.2820 | 0.7543 |
|  | Mixed-race:Complete college | -0.0030 | 0.9970 |
|  | Black:Incomplete postgraduate | -0.0586 | 0.9430 |
|  | Mixed-race:Incomplete postgraduate | -0.1135 | 0.8927 |
|  | Black:Complete postgraduate | 0.6602 | 1.9351 |
|  | Yellow:Complete postgraduate | 0.4494 | 1.5674 |
|  | Mixed-race:Complete postgraduate | 0.1083 | 1.1144 |
| Race/Color (Self-declared) | White | -0.0840 | 0.9194 |
|  | Black | 0.0976 | 1.1025 |
|  | Yellow | 0.0764 | 1.0794 |
|  | Indigenous | -0.2819 | 0.7543 |
|  | Mixed-race | -0.0012 | 0.9988 |
| Main source of income | Student | -0.1733 | 0.8409 |
|  | Self-employed | 0.0135 | 1.0136 |
|  | No source of income | 0.0031 | 1.0031 |
|  | Other | 0.1090 | 1.1152 |
| Race/Color (Self-declared):Main source of income | Black:Student | 0.0930 | 1.0974 |
|  | Mixed-race:Student | -0.3515 | 0.7036 |
|  | Black:Self-employed | 0.1322 | 1.1413 |
|  | Mixed-race:Self-employed | -0.0322 | 0.9683 |
|  | Black:No source of income | -0.2313 | 0.7935 |
|  | Mixed-race:No source of income | 0.0787 | 1.0819 |
|  | Black:Other | 0.1149 | 1.1217 |
|  | Mixed-race:Other | 0.1504 | 1.1623 |
| Predictors or categories without an estimated effect had their coefficients shrunk to zero through penalization and were removed from the table. poly = polynomial transformation (e.g. poly(, 2)1 is the first-degree coefficient of a two-degree polynomial transformation); PR = prevalence ratio. | | | |

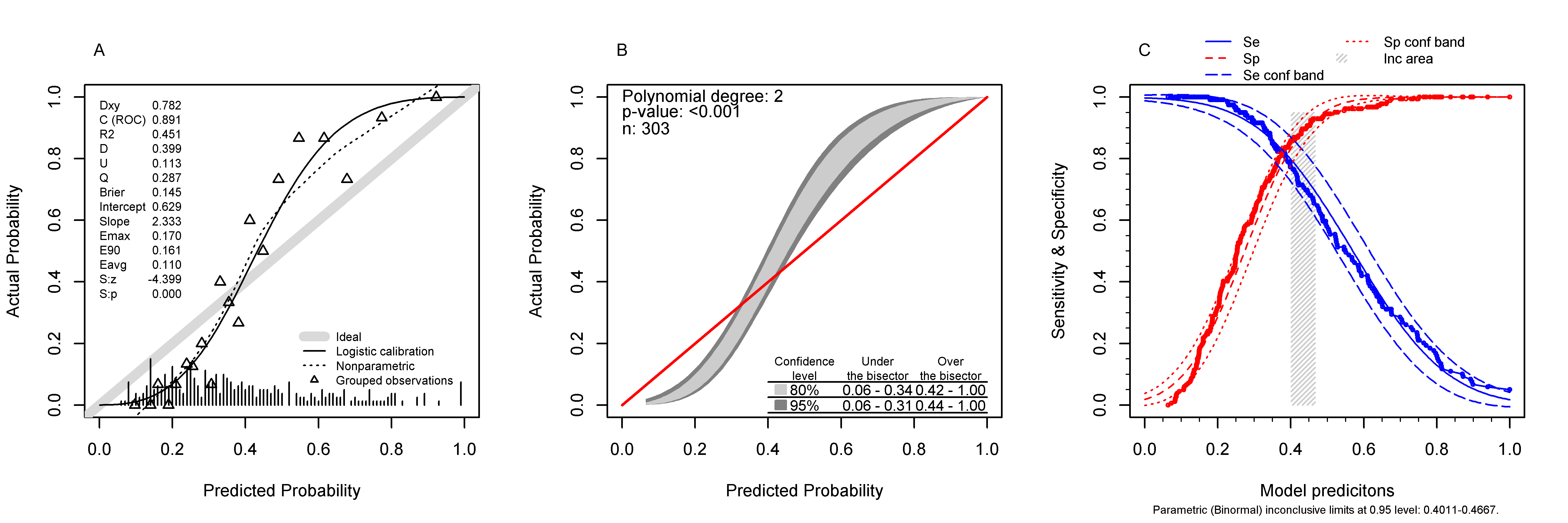

**Figure S****15:** Final penalized and cross-validated Poisson General linear model discrimination and calibration performance without the place of birth and living place as a predictor variable.
